# Altered Toxicological Endpoints in Humans with Quaternary Ammonium Compound Exposure

**DOI:** 10.1101/2020.07.15.20154963

**Authors:** Terry C. Hrubec, Ryan P. Seguin, Libin Xu, Gino A. Cortopassi, Sandipan Datta, Valerie A. McDonald, Claire A. Healy, Najaha A. Musse, Tyler C. Anderson, Richard T. Williams

## Abstract

Humans are extensively exposed to Quaternary Ammonium Compounds (QACs). QACs are ubiquitously used in medical settings, restaurants, and homes as cleaners and disinfectants. They are also used on food and in personal care products as preservatives. Despite their prevalence, nothing is known about the health effects associated with chronic low-level exposure. Chronic QAC toxicity was recently identified in mice and resulted in developmental and reproductive deficits and altered immune function. Cell based studies show that QACs increase inflammation, disrupt cholesterol synthesis, and decrease mitochondria function. If these studies translate to human toxicity, multiple physiological functions could be affected. QAC concentrations in humans have not been monitored previously. This study tested whether QAC concentrations could be detected in the blood of 43 random volunteers, and whether QAC concentrations were associated with markers of inflammation, mitochondrial function, and cholesterol synthesis in a dose dependent manner. QAC concentrations were detected in 80% of study participants, and were associated with decreased mitochondrial function and an increase in inflammatory cytokines in a dose dependent manner. Cholesterol synthesis pathway intermediaries were generally increased, indicating disruption in cholesterol homeostasis. This is the first study to demonstrate that chronic exposure to QACs results in measurable concentrations in human blood, and to also demonstrate significant correlations between QAC level and meaningful biomarkers related to health.

**GRAPHICAL ABSTRACT:** 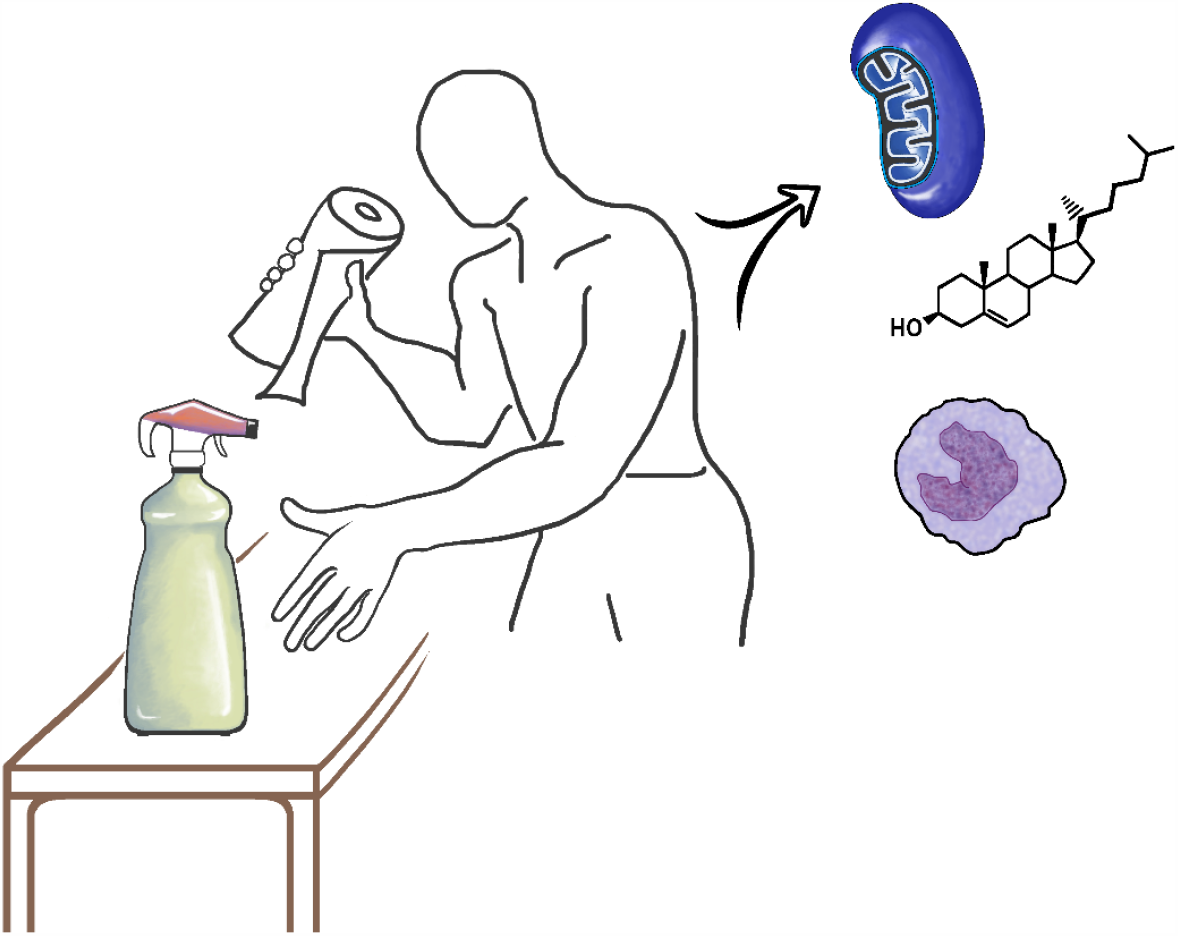

## INTRODUCTION

Humans are extensively exposed to quaternary ammonium compounds (QACs). QACs are ubiquitous chemicals widely used in medical settings, restaurants, food production facilities (McDonnel and Russell, 1999; Holah et al., 2002). They are also used in the home as common cleaners and disinfectants. QACs are frequently added to personal care products and on fresh foods as preservatives. Additionally, QACs, have wide-ranging industrial applications. Despite their prevalence, little is known about the extent of human exposure and the consequences of chronic low-level contact.

Two common QACs are alkyl dimethyl benzyl ammonium chloride (ADBAC, also termed benzalkonium chloride or BAC) and didecyl dimethyl ammonium chloride (DDAC). Over 750,000 tons of both BAC and DDAC are manufactured each year in the US and in Europe (Steichen, 2001). This level of production identifies BAC and DDAC as High Production Volume Chemicals by the US EPA. During production of BAC, a mixture of products is formed. These products contain a benzyl group bound to central nitrogen with alkyl chains composed of differing numbers of carbons units. The average chain length and the distribution of chain lengths vary according to the starting material and the synthetic process used (Danish EPA, 2000). Most processes favor production of chain lengths containing 10 to 18 carbons. This means that BAC solutions contain a mixture of distinct chemicals, each having their own toxicity. Commercial cleaning products often contain mixtures that are predominantly C14 BAC or C12 BAC, followed by C16 BAC. Additionally, many commercial products also contain 2 or more QAC species in combination, such BAC and DDAC. This is important as chemical mixtures can act synergistically or antagonistically to produce an effect that is different from the sum of the individual components. Evaluation of common mixtures is essential in determining chemical exposure risk (Hermens et al., 1985; Faust et al., 2003; Narotsky et al., 2011).

QAC disinfectants are historically viewed as having low toxicity. Accidental overdoses from ingestion of the concentrated mix have resulted in human deaths, but these events are rare (Chataigner et al., 1991; Ellenhorn et al., 1997). Chronic exposure is known to cause asthma and contact dermatitis (Bernstein et al., 1994; Suneja and Belsito, 2008), as well as ocular inflammation and hypersensitivity (Hong and Bielory, 2009). Risk assessments, conducted by the US EPA in 2006, determined that both BAC and DDAC pose little risk to humans (EPA 2006a, 2006b). When the risk assessment is examined in detail, it is clear that the determination was based on studies with decreased body weight as the toxicological endpoint. These studies, using rats and rabbits, found decreased weight gain in adults, neonates, and fetuses at the higher doses (Tyl, 1989; Neeper-Bradley, 1990, 1993). No other specific and subtle toxicological endpoints have been tested. More importantly, no studies have evaluated the extent and magnitude of human exposure, or the possible systemic response to this exposure.

Chronic QAC toxicity has only recently been identified. We have shown using *in-vitro* studies on both human and mouse cell lines that QACs increase inflammation, disrupt mitochondrial function, alter estrogen signaling, and inhibit cholesterol synthesis (McDonald, 2017; Datta et al., 2017; Herron et al., 2016). In mice, we have shown that a commercial cleaner containing a mixture of BAC+DDAC used to disinfect the vivarium, affected reproduction, embryo development, and immune function (Melin et al., 2014, 2015; Hrubec et al., 2017; McDonald, 2017). We have also shown that BACs can cross the placenta and alter cholesterol and lipid homeostasis in mouse neonatal brains after gestational exposure through the maternal diet (Herron et al., 2019). If these studies translate to human toxicity, many basic physiological functions and disease processes could be altered. The body of evidence from our animal and cell based studies suggests toxicity on both a cellular and organismal level. Extrapolation from animal and *in-vitro* studies is the foundation for assessing the safety of drugs and chemicals. Thus, there is a high potential for QAC exposure to cause adverse effects in humans. There is, however, a complete data gap on the effects of chronic low-level systemic exposure to QACs. The objective of this study was to determine the extent of human exposure and determine if there is a systemic response associated with exposure.

## METHODS

Our cell and mouse-based studies suggested that QACs promote increased proinflammatory cytokines, decreased mitochondrial function, and inhibition of cholesterol synthesis. Therefore, we focused on these potential consequences in this current assessment of human QAC exposure.

### Sample Population Characteristics

The blood samples were collected from participants recruited from Blacksburg, VA in the USA. Blacksburg is a small, college town, largely comprised of individuals associated with Virginia Tech, a large university. Blood samples were collected anonymously, and no personal identifying information was collected. Based on visual categorization of age, approximately 2/3^rds^ of the study participants were likely college students, and 1/3^rd^ older adults likely affiliated with the university in some capacity. Inclusion criteria included: non pregnant individuals who were greater than 18 years of age with no history of chronic illness, no acute illness, and previous blood draw in the last two weeks. All research was approved by the IRB (project number 1302366-8) for the protection of human subjects.

### Sample Size Calculation

We hypothesize that there is an association between measured toxicological endpoints and QAC concentration in blood. By estimating the correlation coefficient to be 0.5 and setting α=0.05 and power=0.9, at least 31 blood samples were needed to determine associations between blood analytes and QAC concentrations.

### Sample Collection

Blood samples were collected from 43 healthy volunteers by a certified phlebotomist following IRB approved protocols to protect human subjects. Blood was collected from each participant into 3 separate heparinized blood tubes. One tube was used for mitochondrial analysis, a second tube was used for sterol and QAC analysis, while the third tube was used for cytokine analysis. Determination of each blood analyte was made blind to the participant’s QAC concentration.

### Mitochondrial Function

Mitochondrial physiology was assessed with an XF24 Extracellular Flux Analyzer (Seahorse System, Agilent, Santa Clara, CA) which determines O_2_ consumption under different test conditions. Briefly, 6M white blood cells were aliquoted onto Seahorse XF24 plates at ∼20,000 cells per well. Oxygen consumption rate (OCR) was determined at basal rate (ATP synthesis inhibited by oligomycin), maximal mitochondrial stimulation from addition of FCCP, and at full mitochondrial inhibition from addition of Antimycin A/rotenone (ORC from Proton Leak).

### Sterol Analysis

Analysis of Cholesterol and the cholesterol synthesis precursors: 7-Dehydrocholesterol (7-DHC), 8-Dehydrocholesterol (8-DHC), 7-Dehydrodesmosterol (7-DHD), Lanosterol, Zymosterol, Lathosterol, and Desmosterol, were determined by mass spectrometry. Lipids were extracted from the whole blood and the sterols analyzed by reverse phase LC-MS/MS assays as described previously (Herron et al., 2018).

### Cytokine Production

Cytokines were determined by ELISA. C-reactive protein (CRP) was determined in plasma (R&D Systems, Minneapolis, MN) and NF-κB determined in whole blood (ABNOVA, Taipei, Taiwan). The remaining cytokines, IL-6, IL-10, IL-12, and TNFα, were determined in plasma (R&D Systems) under three conditions. The first determination was an unstimulated baseline prior to culture and stimulation. The remaining whole blood was divided into two aliquots. One received no additives while the other was stimulated with addition of 1µg/mL LPS. Both were cultured simultaneously as a whole blood culture for 12 hours at 37°C with 5% CO_2_. After culture, plasma was collected and assessed for cytokine concentration. There was no difference between the unstimulated baseline and the unstimulated cultured control, thus the unstimulated cultured control was not included in further analysis. IL-10 is generally considered an anti-inflammatory cytokine, but stimulation with LPS alone converts macrophages to the M1 pro-inflammatory phenotype and results in pro-inflammatory IL-10 production.

### QAC Determination

QAC levels in the blood were measured to accurately gauge exposure. Blood samples were spiked with known amounts of deuterium-labeled (*d*_*7*_-benzyl) BACs as internal standards (Herron et al., 2016). All samples were extracted by Folch solution (chloroform/methanol = 2/1). After extraction, the samples were re-constituted in the LC solvent and then analyzed by UPLC-MS/MS (Seguin et al., 2019).

### QAC Nomenclature

The nomenclature used to describe QACs varies across publications due to the large number of chemicals in the QAC class, as well as the fact that many common QACs have multiple synonyms. For the purposes of this paper, the following nomenclature will be used. QAC is used to refer to both BAC and DDAC together. BAC, with no modifier, is used when speaking of the various BAC alkyl chain lengths together, while the modifiers C10, C12, C14 and C16 BAC are used to designate the specific BAC chemical with the indicated alkyl chain length.

### Statistical Analysis

Univariate descriptive statistics were conducted. QAC concentrations were then compared with absolute values of mitochondrial function, sterol synthesis intermediaries, and cytokine production to determine associations between the toxicological endpoints and the blood QAC concentration. As both the toxicological endpoints and QAC concentration measurements are continuous, the nonparametric Spearman’s Rank Correlation (r_s_) was used to measure associations between all endpoints. Each QAC species was ranked high to low and correlated with the associated analyte value for that sample. Associations were calculated across the ranked samples, increasing by 1 sample (N) in each calculation. For example, 10 individuals contained C12 BAC in their blood. The 10 samples were ranked from high to low. Correlations were determined between C12 BAC and each analyte for the five individuals with the highest C12 BAC. A second correlation was then calculated for individuals with the six highest C12 BAC concentration. The process was repeated until correlations were determined for all 10 individuals containing C12 BAC. Correlation coefficients of 0.5 with a p < 0.05 were used to identify significance. This is an accepted level of association in studies relating concentrations of an environmental contaminant with a pathophysiological outcome (Li et al., 2011; Jenkins et al., 2002; Bárány et al., 2002). While this correlation coefficient may seem low compared to a tightly controlled laboratory study, the normal variability in blood constituents, and variations in exposure level from randomly sampled individuals, necessitates a more relaxed measure of significant association. For means comparison tests, each blood analyte was stratified into two groups for each QAC; samples with no QAC and samples with QAC. Difference in the means of the two groups was determined by a t-test for parametric data, or Wilcoxin Rank Sum Test for non-parametric data. Each blood analyte was also stratified into low, medium and high evenly spaced QAC terciles. Differences between the means of each tercile were compared by ANOVA for parametric data, or Kruskal Wallis test for non-parametric data. Nineteen of the twenty-seven analytes measured (including QACs) exhibited outliers (determined by Statistix Software, Talahasee, FL). Typically only a single high outlier was seen per analyte; four analytes had two outliers. All outliers were greater than two standard deviations from the mean; 73% of identified outliers were greater than three standard deviations from the mean. The identified outliers were not included in the analysis.

## RESULTS

Approximately 80% of individuals contained measurable concentrations of QACs in their blood. Blood concentrations of each QAC species are given in supplementary data (Table S1). One-third of individuals had Total QAC concentrations between 10 – 150 nM, which has been shown to have physiological effects in cell culture models (Datta et el., 2017; Herron et al., 2016). Descriptive statistics for the complete dataset are given in Table S1. Data were analyzed in two ways; by means comparison, and by association with each QAC. The means comparison test evaluates differences between concentration groups rather than at the individual level, and includes individuals with no QAC residues. The association test measures the direct linear relationship between two variables from each individual across the sample population. Thus, the association test evaluates data at the individual level in relation to the whole dataset, and does not include individuals without QACs in their blood.

### Means Comparison

For analysis, the measured blood concentrations were stratified into the following exposure groups: no QAC, and then low, mid, and high evenly spaced QAC tercile groups. Exceptions were made for C10 and C12 BAC. Only a single individual contained detectable C10 BAC and no further analysis was conducted for this BAC. Furthermore, as only a limited number of individuals contained C12 BAC residues, this BAC was divided into no C12, low C12, and high C12 groups with no mid-range group. Means were then compared between the groups to identify if a dose response was present.

Eleven individuals had C12 BAC residues in their blood; one individual’s value was identified as an outlier (6.5 standard deviations from the mean) and was not included in the analysis. For C12 BAC, significant differences in means were observed for Baseline IL-12 and Stimulated IL-10 (Fig. 1). Both analytes were significantly elevated in individuals with the higher concentrations of C12 BAC in their blood.

**Figure 1.**
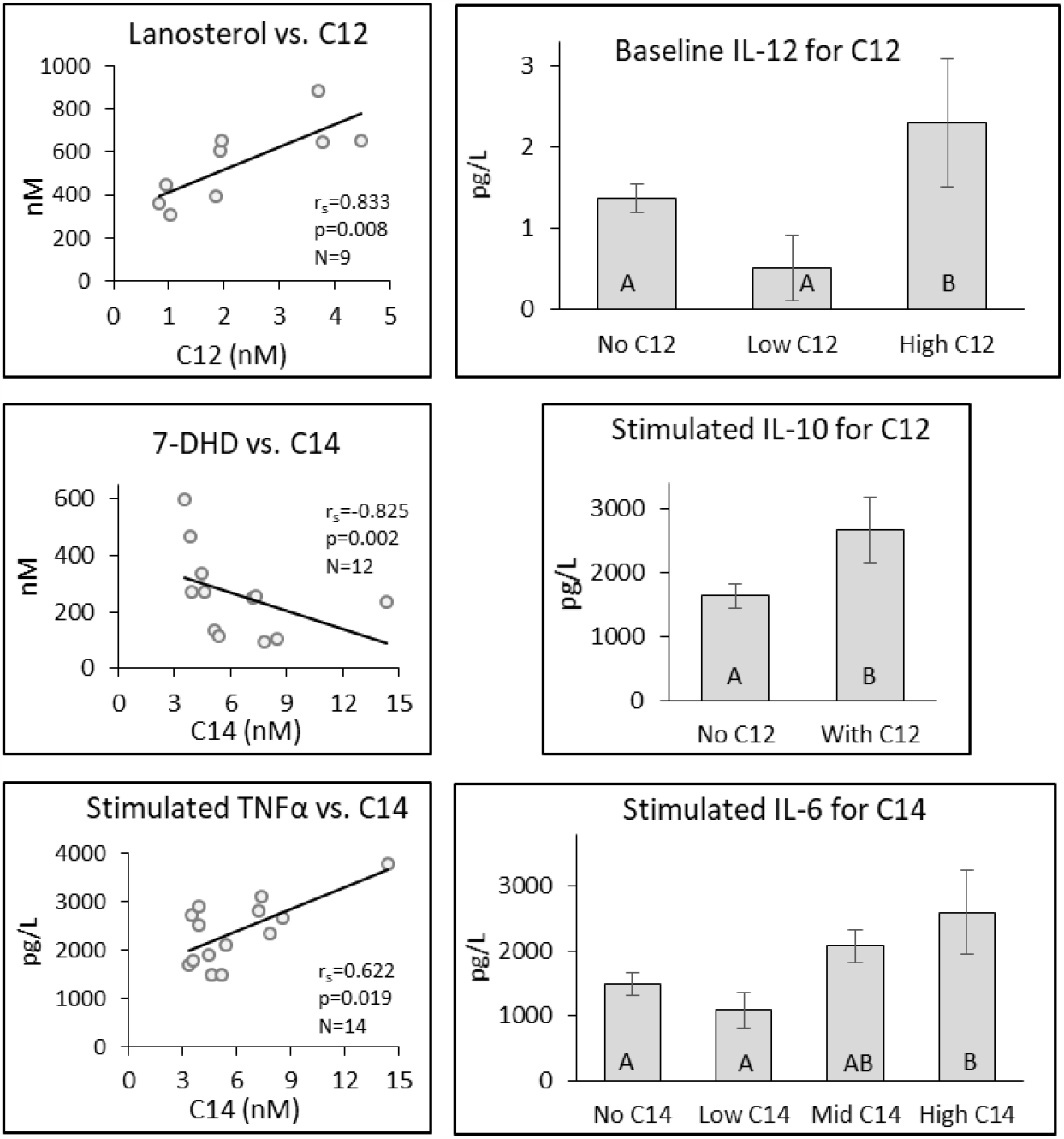
Differences in selected analytes by C12 and C14 BAC concentration found in the blood. Individual bar graphs depict the mean ± SE for different analytes. Bars identified by different letters indicate a significant difference (p ≤ 0.05) between groups. Analytes stratified into two groups were compared by either t-test or Wilcoxin Rank Sum Test, while those stratified into four groups were compared by ANOVA or Kruskal Wallis test. Scatterplot correlations were determined by Spearman’s Rank Correlation (r_s_).

Twenty individuals contained C14 BAC residues in their blood; two individual’s values were identified as outliers (3.8 and 5.5 standard deviations from the mean) and were not included in the analysis. C14 BAC affected IL-6 a common marker of inflammation demonstrating a dose response across the tercile groups (Fig. 1).

Twenty-four individuals contained C16 BAC residues in their blood; one individual’s value was identified as an outlier (6.3 standard deviations from the mean) and was not included in the analysis. C16 BAC affected sterol homeostasis, inflammation, and mitochondrial function (Fig. 2). Individuals with C16 BAC residues in their blood demonstrated an inverted-U nonmonotonic dose response for the cholesterol synthesis intermediary 8-DHC with the highest concentrations found at the low and mid C16 terciles (Fig. 2). The inflammatory markers, Stimulated IL-6 and Stimulated IL-10, demonstrated a J-shaped nonmonotonic dose response with significantly lower blood concentrations in the low and mid C16 BAC terciles. The mitochondrial respiration marker, Maximum OCR from Basal, generally decreased across the exposure groups. Mitochondrial function also demonstrated a significant decrease in individuals with C16 BAC exposure compared to those with C16 BAC in their blood (Fig. 2).

**Figure 2.**
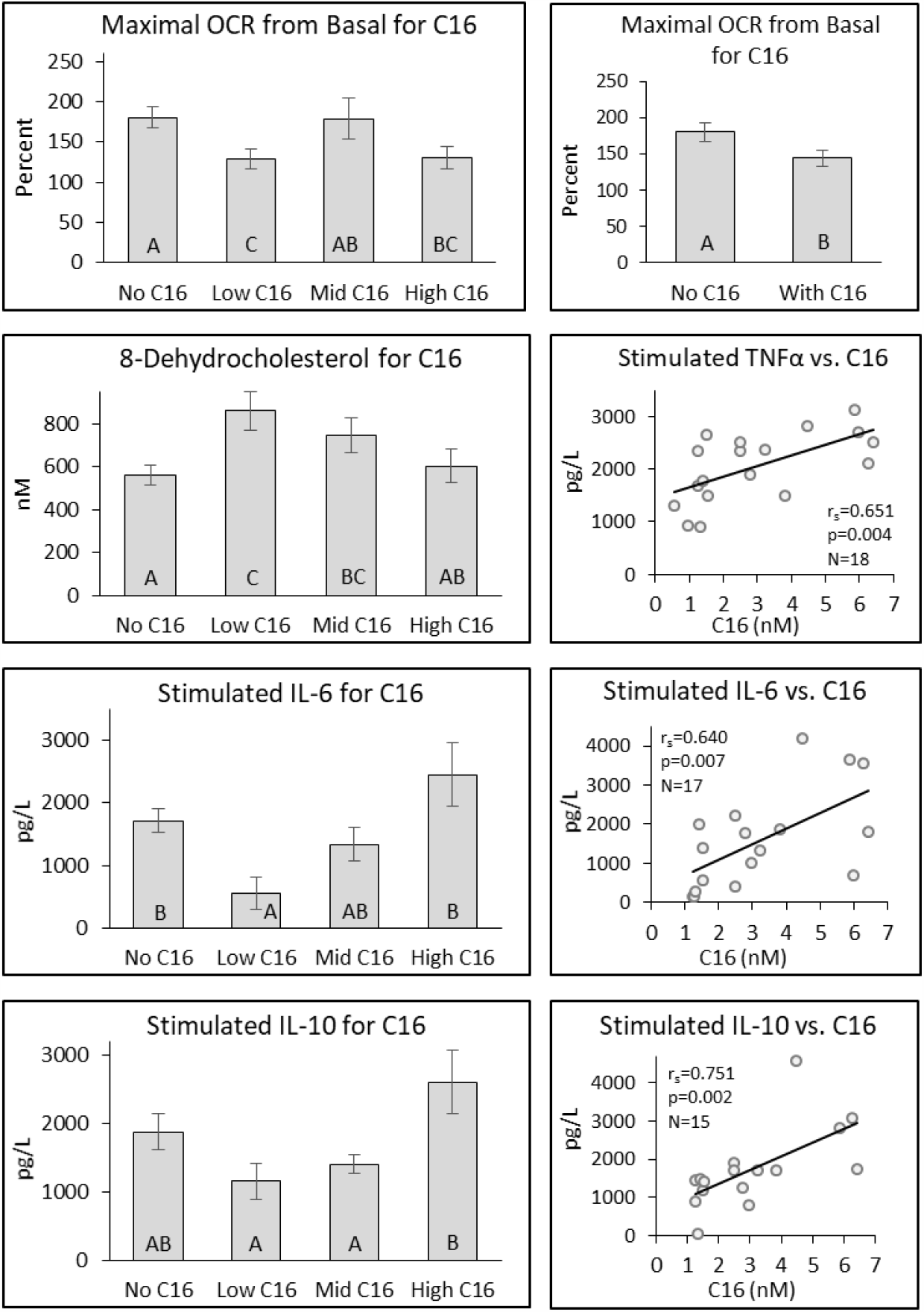
Differences in selected analytes by C16 BAC concentration found in the blood. Individual bar graphs depict the mean ± SE for different analytes. Bars identified by different letters indicate a significant difference (p ≤ 0.05) between groups. Analytes stratified into two groups were compared by either t-test or Wilcoxin Rank Sum Test, while those stratified into four groups were compared by ANOVA or Kruskal Wallis test. Scatterplot correlations were determined by Spearman’s Rank Correlation (r_s_).

DDAC residues were seen in 32 of the 43 participants; one individual’s value was identified as an outlier (5.9 standard deviations from the mean) and was not included in the analysis. DDAC affected cholesterol biosynthetic pathway intermediates, inflammatory markers, and mitochondrial function (Fig. 3). 8-DHC exhibited a difference between individuals with DDAC in their blood compared to those with no DDAC exposure. Baseline TNFα was more than an order of magnitude higher in individuals with DDAC present in their blood than those without DDAC. Baseline IL-10 and Stimulated IL-6 both exhibited a J-shaped nonmonotonic dose response with the lowest IL-10 concentrations found in the low and mid DDAC groups.

**Figure 3.**
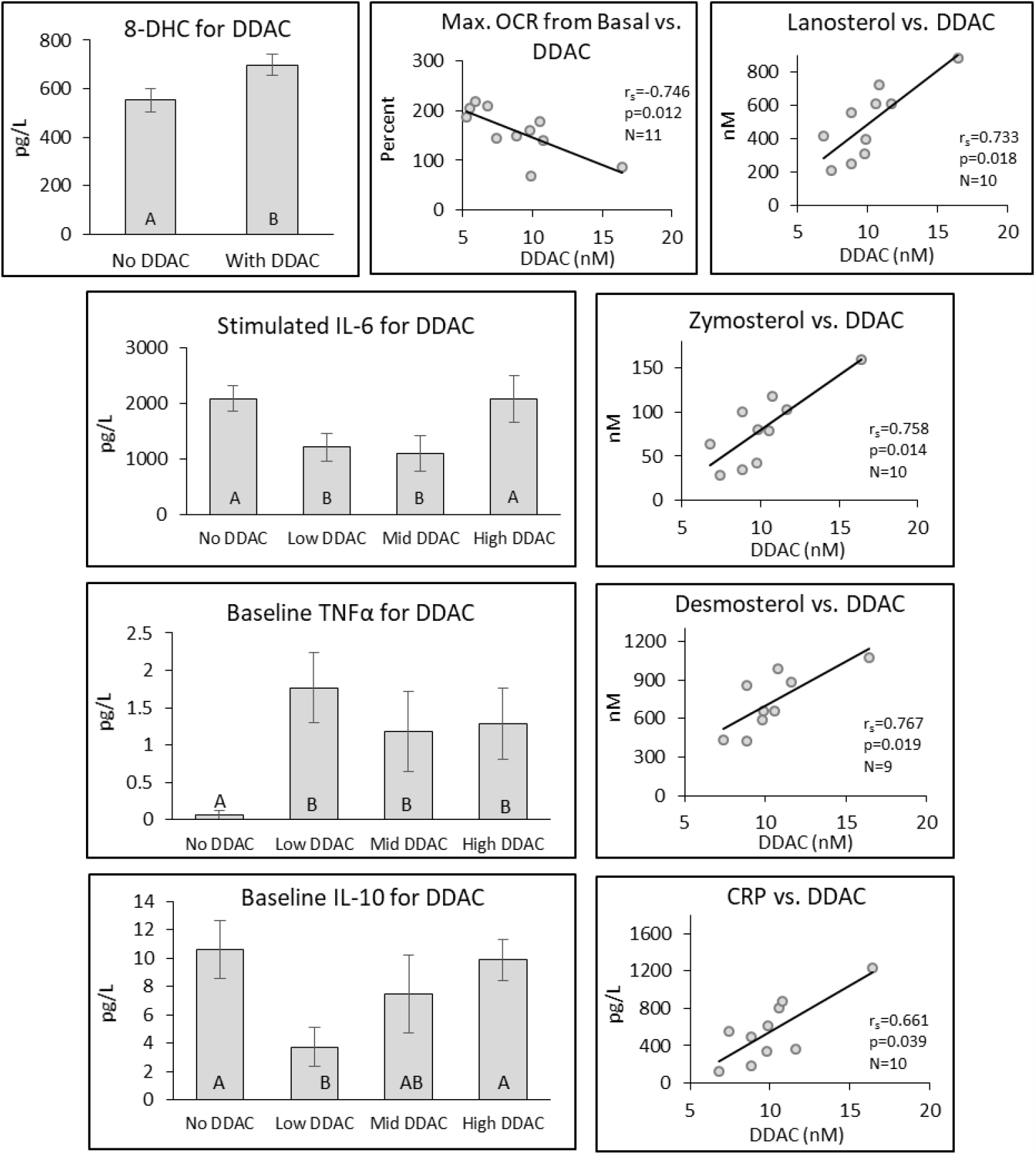
Differences in selected analytes by DDAC concentration found in the blood. Individual bar graphs depict the mean ± SE for different analytes. Bars identified by different letters indicate a significant difference (p ≤ 0.05) between groups. Analytes stratified into two groups were compared by either t-test or Wilcoxin Rank Sum Test, while those stratified into four groups were compared by ANOVA or Kruskal Wallis test. Scatterplot correlations were determined by Spearman’s Rank Correlation (r_s_).

Among our sample population, 35 of the 43 individuals contained QAC in their blood; two values were identified as outliers (2.9 and 5.9 standard deviations from the mean) and were not included in the analysis. When the analytes were evaluated in relation to the Total QAC concentration, the responses were similar to those seen for the individual QAC that had the greatest influence on blood analyte (Fig. 4). This is not surprising as the Total QAC determination is just the sum of all 4 QAC species of meaningful exposure (C12, C14, C16 BACs, and DDAC). Stimulated IL-6 showed a dose response similar to that seen with C14 BAC, C16 BAC, and DDAC. An additive response was seen across the QAC species for the means comparison test of Maximal ORC from Basal. Mitochondrial respiration was decreased at higher concentrations of both C16 BAC and DDAC, however a greater decline in respiration was seen when the two QACs were combined in Total QAC analysis. A cancellation of effect was seen with 8-DHC. 8-DHC concentrations were increased with both C16 BAC and DDAC, but with Total QAC. This cancellation of effect is likely due to the modest declines in 8-DHC concentrations with C-12 and C14 BAC blunting the overall response.

**Figure 4.**
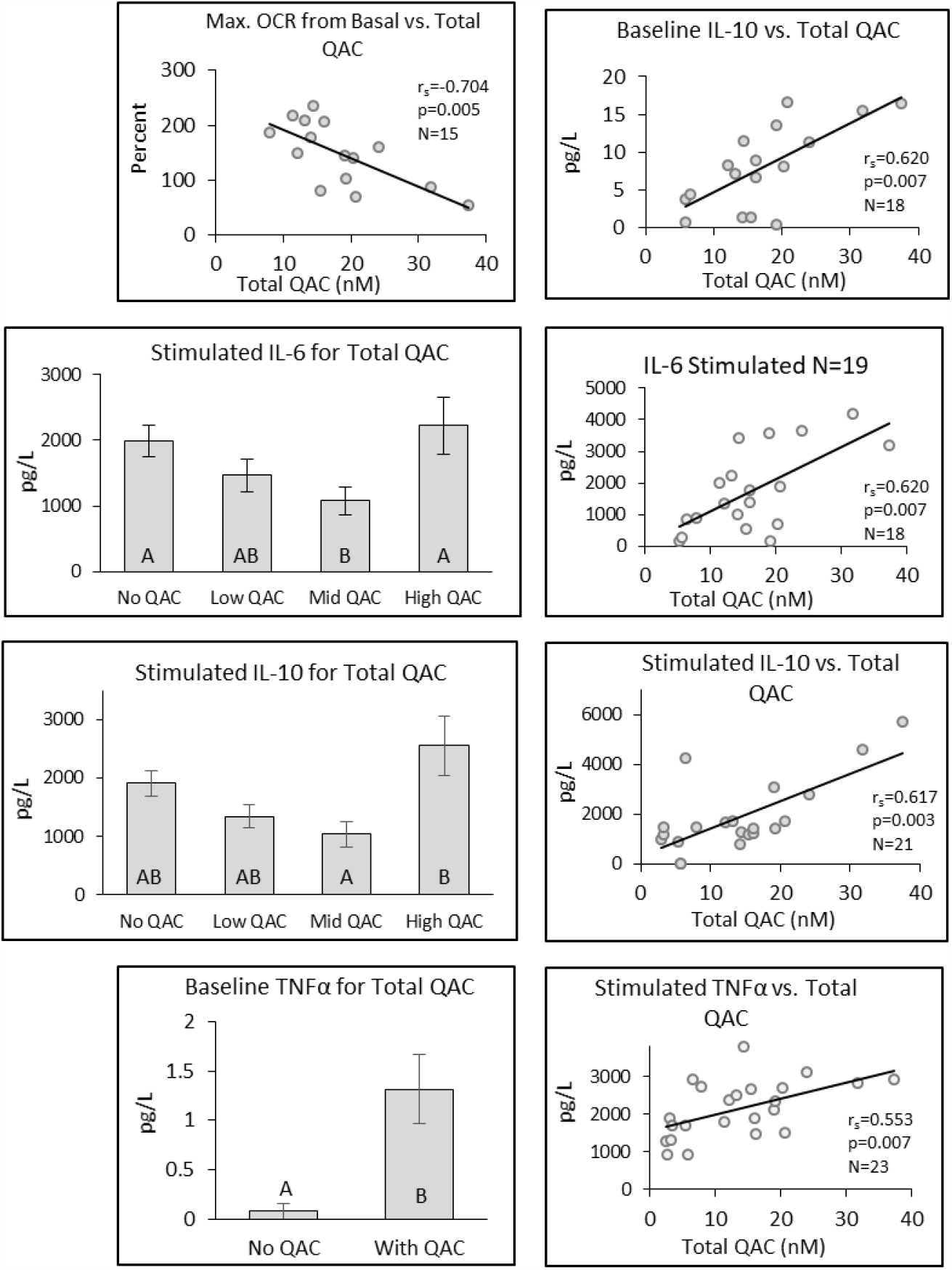
Differences in selected analytes by Total QAC concentration found in the blood. Individual bar graphs depict the mean ± SE for different analytes. Bars identified by different letters indicate a significant difference (p ≤ 0.05) between groups. Analytes stratified into two groups were compared by either t-test or Wilcoxin Rank Sum Test, while those stratified into four groups were compared by ANOVA or Kruskal Wallis test. Scatterplot correlations were determined by Spearman’s Rank Correlation (r_s_).

### Associations

Significant associations (r_s_) were observed between a number of analytes and specific QAC species. As expected, the associations varied with the number of samples included in the calculation. All calculated associations are given in Supplemental Tables 2 - 6.

For C12 BAC, the cholesterol pathway constituent Lanosterol demonstrated a strong positive correlation (r_s_=0.833, p=0.008, N=9) with C12 concentrations between 4.468 and 0.828 nM across 9 of the 10 individuals (Figs. 1 & 5). The increase in the precursor Lanosterol indicates a slowdown in a downstream step of the cholesterol synthesis pathway or a general upregulation of cholesterol production.

**Figure 5.**
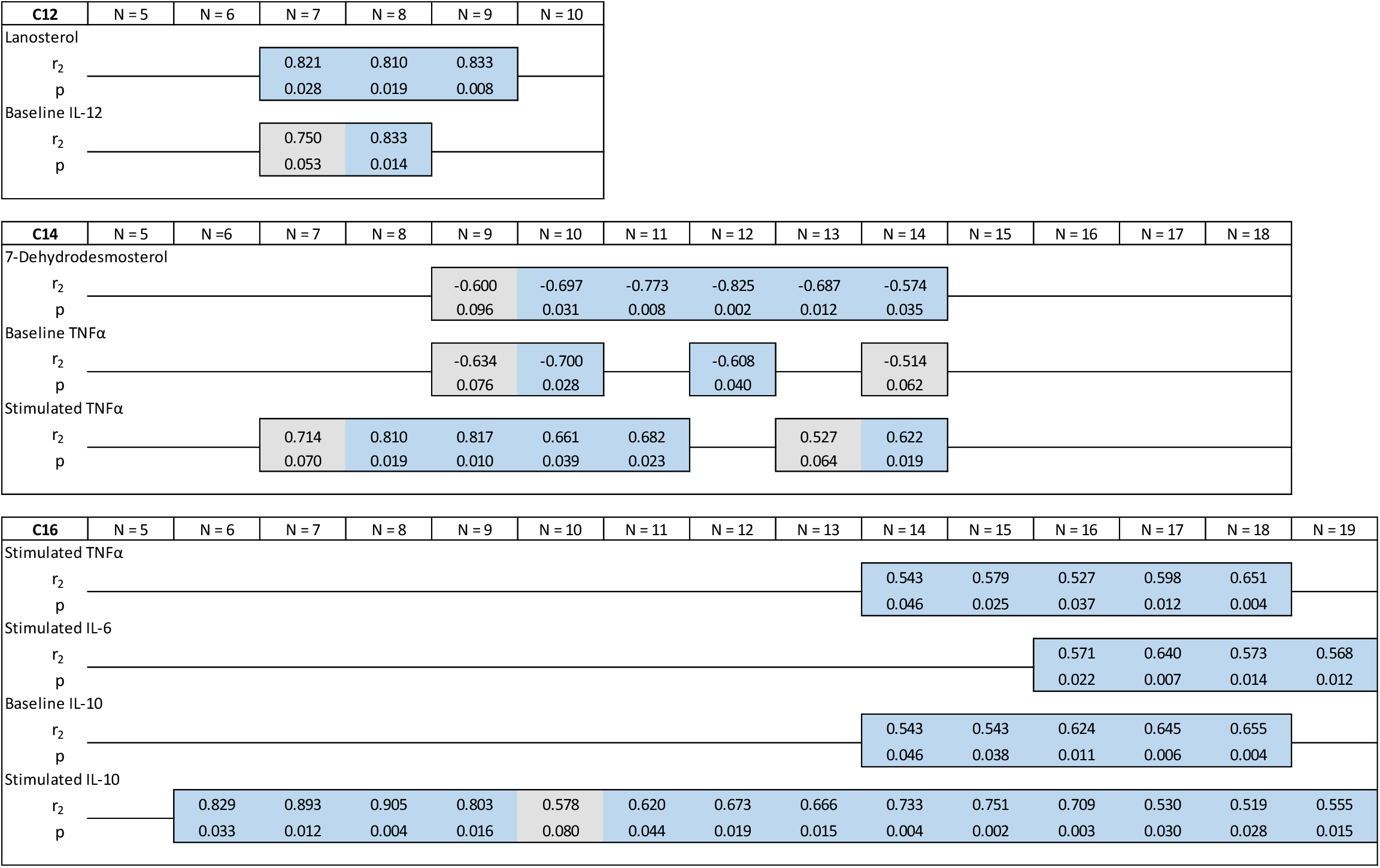
Significant associations of C12, C14, and C16 BAC with blood analytes. The number of individuals with C12, C14, and C16 BAC detected in their blood was 10, 18, and 22 respectively. Each QAC species was ranked high to low and correlated with the associated analyte value for that sample. The N is the number of ranked samples included in the calculation. Correlation r_s_ values were determined by the nonparametric Spearman’s Rank Correlation test. Correlation coefficients ≥ 0.5 with a p ≤ 0.05 are shaded blue and considered significant. Correlation coefficients ≥ 0.5 with a p ≤ 0.1 are shaded grey to designate associations with less certainty. Significant correlations were not found beyond an N of 19 for C16 BAC and are not shown.

For C14 BAC, both cholesterol synthesis intermediates and inflammatory markers demonstrated associations (Figs. 1 & 5). 7-DHD demonstrated a strong negative association (r_s_=-0.825, p=0.002, N=12) which remained significant over a range of N=10 to N=14 (from 14.383 to 3.337 nM C14 BAC) and with less certainty for N=9. Baseline TNFα was significantly correlated (r_s_=-0.700, p=0.028, N=10) with C14 BAC; however, the association was inconsistently present. On the other hand, Stimulated TNFα was strongly and significantly associated with C14 BAC across much of N=8 to N=14 (from 14.383 to 3.337 nM C14 BAC) with the highest association seen at r_s_=0.817, p=0.010, N=9 (Fig. 1 & 5).

Associations were observed with C16 BAC only for inflammatory intermediaries (Fig. 2 & 5). Stimulated TNFα, and Stimulated IL-6 were weakly associated with C16 BAC (r_s_=0.640, p=007, N=17; and r_s_=0.563, p=0.013, N=19, respectively). The association was significant from C16 BAC concentrations of 6.411 to 0.935 nM. On the other hand IL-10, in both Stimulated and Baseline samples, was positively associated with C16 BAC. Stimulated IL-10 showed strong correlations across the sample population from N=6 to N=19. The strongest correlation was seen with N=8 (r_s_=0.905, p=0.004) but significant correlations continued through N=19 (from 6.411 to 0.935 nM C16 BAC). Baseline IL-10 was more weakly correlated (r_s_=0.655, p=0.004). While usually considered an anti-inflammatory cytokine, IL-10 has both pro and anti-inflammatory properties. Stimulation with LPS alone as was done in this study, converts macrophages to the M1 pro-inflammatory phenotype and results in pro-inflammatory IL-10 production. It is not clear whether IL-10 production in the Baseline sample represented a pro-inflammatory or anti-inflammatory stimulus.

Strong positive correlations were found with DDAC and a number of cholesterol synthesis pathway intermediaries including: Cholesterol, Zymosterol, Desmosterol, 7-DHD, and Lanosterol (Fig. 3, Fig. 6). For Cholesterol, the strongest correlation (r_s_=0.893, p=0.012) was seen with N=7, but significant correlations were seen from N=6 to N=9 (from 16.421 to 7.432 nM). For Zymosterol, the strongest correlation (r_s_=0.929, p=0.006) was seen with N=7, but significant correlations were seen from N=6 to N=13 (from 16.421 to 5.940 nM). For Desmosterol, the strongest correlation (r_s_=0.929, p=0.006) was seen with N=7, with significant correlations seen from N=6 to N=9. Associations with less certainty were seen to N=13 (from 16.421 to 7.432 nM). For Lanosterol, the strongest correlation (r_s_=0.893, p=0.012) was seen with N=7, but significant correlations were seen from N=6 to N=11 (from 16.421 to 6.479 nM). In addition to cholesterol synthesis pathway intermediaries, Maximal Mitochondrial OCR from Basal showed strong negative correlations with DDAC (r_s_=-0.745, p=0.012) to N=11 (from 16.421 to 6.479 nM) (Figs. 3, Fig. 6, Table S5). Additionally, the inflammatory markers CRP and Stimulated IL-10 were positively associated with DDAC concentration. Thus, while DDAC had the greatest influence on the cholesterol synthesis pathway, mitochondrial and inflammatory markers were affected as well.

**Figure 6.**
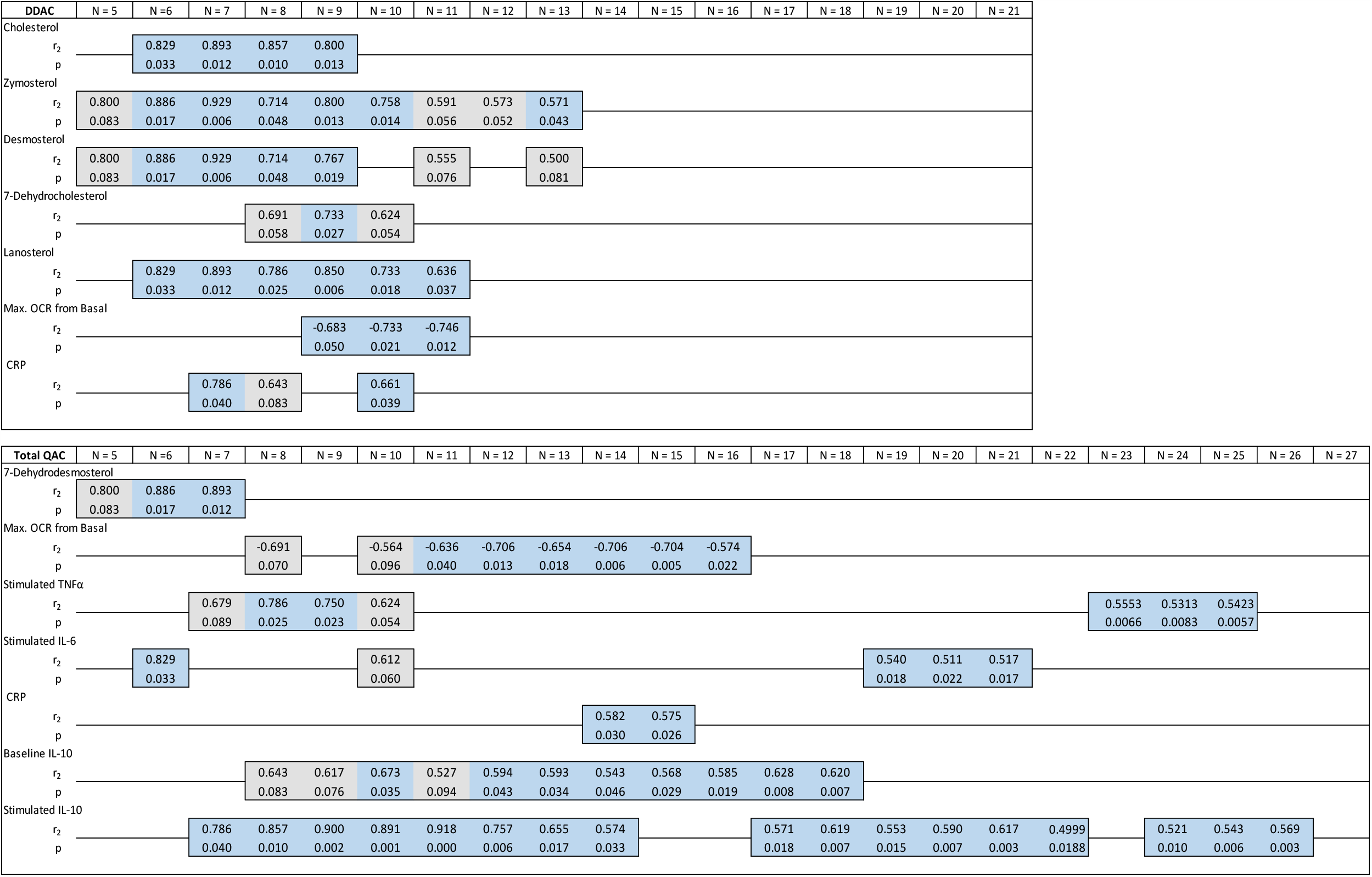
Significant associations of DDAC and Total QAC with blood analytes. The number of individuals with DDAC, and with the sum of all QAC species (Total QAC) in their blood was 31 and 33 respectively. Each QAC species was ranked high to low and correlated with the associated analyte value for that sample. The N is the number of ranked samples included in the calculation. Correlation values were determined by the nonparametric Spearman’s Rank Correlation test. Correlation coefficients ≥ 0.5 with a p ≤ 0.05 are shaded blue and are considered significant. Correlation coefficients ≥ 0.5 with a p ≤ 0.1 are shaded grey to designate associations with less certainty. Significant correlations were not found beyond an N of 20 for DDAC and 26 for Total QAC, and are not shown.

Total QAC concentration represents the sum of the 4 individual constituents C12, C14, C16 BACs, and DDAC. Associations of analytes in relation to Total QAC concentration can be enhanced from what is seen with each QAC species if the effects are additive, or potentiated. On the other hand, associations can be diminished if there are divergent responses between individual QAC species, or if a response to a single QAC is diluted by a lack of response to the other QAC components. Increased Total QAC concentrations were strongly and significantly associated (Figs. 4 & 6) with decreased maximal mitochondrial function (r_s_=-0.706, p=0.006) from N=11 to N=16. Less certain associations were seen from N=8, over a total QAC range from 37.4 to 5.7 nM. While mitochondrial function was decreased with individual QAC species, the greatest inhibition was seen with the Total QAC comparison. This indicates that the effect in the QAC mixture is likely additive. Strong significant associations were also observed for both Baseline and Stimulated IL-10 production (Fig. 4 &6). These significant associations were seen over much of the sample population. While much of this effect came from the response to C16 BAC, the other BAC species were also positively associated with IL-10 production (see supplemental data) resulting in an additive response to Total QAC. The strong associations between C16 BAC and both TNFα, and IL-6 were not as prominent in the Total QAC mixture. This is most likely due to an obscuring of these responses when Total QACs were assessed as a whole.

## DISCUSSION

While contradictory evidence is mounting, the widely held belief is that although exposure to BAC and DDAC occurs, little if any accumulates systemically as these chemicals were assumed to pass through the intestine largely unabsorbed. This belief is based on unpublished rodent studies conducted for regulatory purposes (Henderson, 1992; US EPA 2006a, 2006b). For BAC, 90-98% of an orally administered dose was eliminated in the feces, and only 0.03 to 0.5% of the dose was retained in body tissues (US EPA 2006a). For DDAC, 89-99% of an oral dose was eliminated, unabsorbed, in the feces and only 0.003-0.675% of the DDAC dose remained in the tissue (Henderson 1992). However, these studies DDAC and BAC rely upon a single oral dose; tissue accumulation following chronic exposure, has not been assessed.. Furthermore, substantial recovery of QAC in feces does not definitively demonstrate low absorption. For example, approximately 1/3^rd^ of excreted BAC and 40% of excreted DDAC are oxidized metabolites (EPA 2006a; EU, Directive 98/8/EC, 2015); and we have recently shown that BAC is converted to oxidized metabolites by hepatic cytochrome P450 enzymes expressed in the liver (Seguin, et al. 2019). The presence of the oxidized metabolites in the feces suggests orally administered BAC and DDAC were absorbed and reached the liver. Others have also determined that the major excretory route of high molecular weight QACs such C12, C14, and C16 BACs, and DDAC is via bile into feces (Hughes, et al. 1973; Smit, et al. 1998; Song, et al. 2013). Thus, the extent of oxidized QAC metabolites in the feces could in fact reflect a greater than appreciated extent of intestinal absorption, followed by hepatic metabolism and biliary excretion back into feces.

QAC exposure can also occur by other routes. Xue et al. (2004) found much higher blood and tissue concentrations when BAC was aspirated into the lungs, or given parenterally than when dosed orally. This is consistent with the efficient metabolism of BACs by liver cytochrome P450 enzymes following an oral dose, as aspiration or parenterally administration would not undergo hepatic first-pass metabolism. We have observed the same pattern of effect in our reproductive and developmental toxicity studies in terms of routes of exposure. Undosed mice that received ambient exposure (most likely inhalant) from use of QAC disinfectants in the vivarium, showed similar degrees of reproductive and developmental impairment as mice receiving both oral and ambient exposure (Melin et al., 2015; Hrubec et al., 2017). This implies that most of the toxicity results from the ambient/inhalant exposure. This is troubling as ambient exposure more closely mimics human exposure from use of cleaning products. There are virtually no studies investigating the systemic toxicity from inhalant exposure of BAC or DDAC. A single study in rats, evaluating inhalant toxicity of cetylpyridinium chloride, a common but structurally unrelated QAC, found mortality after a single exposure in all dose groups including the lowest dose of 50 mg dust/m^3^ (Lin et al., 1991). Calculations of the LD_50_ indicated that systemic toxicity was greater with the inhalant route than other routes. Taken together, these studies show that respiratory tract exposure can result in higher blood concentrations and increased toxicity than oral dosing. Clearly, the human population is exposed, and equally clearly, our results demonstrate that QACs can be absorbed systemically and be detected in human blood, but the main route or routes of exposure are not known.

Approximately 80% of participants in our study had detectable concentrations of BAC and DDAC in their blood; and some individuals contained high concentrations between 10 – 150 nM. The primary source of exposure responsible for the blood residues is not known as QACs are used occupationally as well as in home and public settings. Occupational exposure is an obvious candidate for primary exposure, but daily cleaning of the home in poorly ventilated spaces may result in equivalent uptake of QACs. Additionally, toxicokinetic studies have not been conducted in humans; and thus, the distribution of QACs in the body, rate of metabolism, and eventual clearance is not known. The BAC and DDAC detected in our study population could indicate prior exposure that day, or be the result of low level exposure over months. In 2006, the US EPA risk assessment identified dermal exposure to the concentrated disinfectant, and inhalant exposure to the working concentration of disinfectant as posing the greatest risk to people. This was based on reports of dermal burns for the former and reports of asthma, allergy and hypersensitivity for the latter. The risk assessment did not evaluate internal systemic exposure as it was presumed that BAC and DDAC do not accumulate in the body. Identifying the main source and the route of systemic exposure is critical in order to modify or limit one’s internal dose.

The different QAC species were detected in the following abundance: DDAC > C14 BAC > C16 BAC > C12 BAC > C10 BAC. This roughly follows the proportion of QACs found in most commercial disinfection products; DDAC is usually twice the concentration of the total BAC component, and C12 BAC is frequently double the concentration of C16 BAC. In our blood samples, the average DDAC concentration was only slightly greater than C14 BAC, and C12 BAC was roughly the same concentration as C16 BAC. Single individuals did not necessarily contain all QAC species. Only one individual contained detectable C10 BAC and only 11 individuals contained C12 BAC. DDAC was the most commonly found QAC and was present in 32 individuals. This difference in QAC species distribution indicates either differences in absorption or elimination rates, or that the blood concentrations reflect exposure to multiple formulations and different sources of QAC components in the environment.

While most individuals clustered within a certain range of QAC concentrations, some individuals had a value that was up to an order of magnitude higher than the next closest value. These extreme measurements were not attributable to a single individual, but were found in different individuals. Corresponding measured analytes in these extreme samples were frequently divergent as well. In all cases, neither the QAC outlier nor the associated analyte were included in the statistical analysis. The numbers of sample at these extreme concentrations of QACs were too few to assess a valid response.

Increasing QAC concentrations accompanied significant increases in inflammatory mediators, decreases to mitochondrial function, and decreases in cholesterol synthesis. Analyte response varied with the different alkyl chain lengths of BAC. It is generally held that longer chain QACs have lower toxicity than shorter alkyl chain QACs due to decreased water solubility (Danish EPA, 2000). However, this does not match our previous observations nor the findings of this study. We found that C16 BAC affected more constituents than the other BAC species, despite the fact that C16 BAC was found at lower concentrations in the blood. Overall C16 BAC had the greatest effect on markers of inflammation while DDAC predominantly affected cholesterol synthesis. Mitochondrial function was decreased both with C16 BAC and with DDAC, but demonstrated the strongest response with Total QAC indicating the response was likely additive across the QAC species tested.

In some instances, the means comparison and the association tests gave different results. This is not unreasonable as the two tests measure different aspects of effect. Association tests can tell the concentration range over which there is a potential effect. Ideally associations should be determined on sufficient numbers of individuals in the mid to high exposure range. When only a few individuals at the higher dose range are evaluated, there are too few individuals to establish a true relationship. Conversely, when sufficient individuals at the lowest blood concentrations are included, associations disappear as the QAC concentration falls below the no-observed-effect-level (NOEL). The NOEL indicates the concentration below which the measured QAC has no discernable effect on the particular blood analyte. Chemicals with higher NOELs are less toxic then ones with lower NOELs. From our data, it appears that DDAC has a higher NOEL as across the board the correlations dropped off sooner than for the BAC species.

Means comparison tests can show a dose response across dose groups. Nonmonotonic or hormetic responses are by definition not linear, and thus will not give a linear association across the entire concentration range. This could result in statistically significant associations across only a portion of sample population as was seen for a number of the analytes measured in this study. The idea of a toxicant having a nonmonotonic /hormetic dose response is not new, and is reviewed in detail by Calabrese (2005). First coined in the 1940s, hormetic dose responses demonstrate either an inverted U-shaped curve with decreased responses at low and high doses, or a J-shaped curve with enhanced responses at low and high doses. In an analysis of the published toxicological response literature, Calabrese and Baldwin (2001) estimated that the overall frequency of hormetic dose response across the spectrum of known toxicants is approximately 40%. This includes responses to many well recognized toxicants such as DDT, mercuric chloride, lead, cadmium, chromium, toluene, and others (Calabrese 2005). With regards to QACs, an early study evaluating immunologic effects of BAC, found that histamine release from mast cells followed a nonlinear dose response (Danish EPA, 2000). Additionally, BAC binds to the acetycholine muscarinic receptor, and also the estrogen receptor (Takeuchi et al., 1993; Datta et al., 2017). Many receptor mediated signaling systems, including those involving the muscarinic, and estrogen receptors, have a biphasic nonmomotonic dose response (Calabrese 2005).

Regardless of whether the response is linear or nonmonotonic, our data clearly demonstrated a dose dependent association between blood analytes and the amount of QAC present in the blood. Cytokine production appeared heavily impacted with increases in both baseline and stimulated responses at higher QAC concentrations. Baseline measurements represent the resting/current concentration, while stimulated samples represent the capacity to respond after appropriate signals. The inflammatory effects were particularly striking for TNFα, IL-6 and IL-10. Mitochondrial function was also strongly and significantly inhibited in a dose dependent manner. These changes are similar to those found in our cell and mouse studies. Also corroborating our previous studies, we found that cholesterol synthesis intermediaries were profoundly affected. Increasing concentrations of DDAC were significantly associated with increasing Zymosterol, 7-Dehydrocholesterol, Desmosterol, Lanosterol, and cholesterol while C12 BAC was positively associated with Lanosterol. Previously we demonstrated *in-vitro* that BAC inhibition of the final step of cholesterol synthesis led to upstream accumulation of the synthesis precursors, particularly 7-DHC and 8-DHC (Herron et al., 2016). However, results in the present study suggest that there is an overall upregulation of cholesterol biosynthesis by DDAC. Interestingly, as C14 BAC increased, 7-DHD decreased indicating that C14 BAC may act directly to inhibit conversion of Zymosterol to 7-DHD. These results suggest that different types of QACs may have different effects on cholesterol biosynthesis.

The correlations found in the present study are strong. The accepted significant correlation coefficient for evaluating environmental effects in human tissues is 0.5 (Li et al., 2011; Jenkins et al., 2002; Bárány et al., 2002). We observed significant correlations from 0.5 to 0.9 for a number of the blood analytes measured. It must be kept in mind however that this is a small pilot study with a limited sample size. No personal information about the study population was collected. As such, confounding influence of external variables such as diet, weight, exercise status, or pharmaceutical use of cholesterol modulating drugs is not known. Even with this caveat, these data are provocative and indicate possible ramifications for health and disease. If the changes observed in this study translate to the population at large, and are associated with pathophysiology, then QAC exposure could be involved in many and varied disease processes. Potentially impacted may be inflammatory diseases such as diabetes, obesity, osteoarthritis, autoimmunity, and cancer; mitochondrial associated diseases such as cardiovascular disease, obesity, and neurodegenerative disease. Disruptions to lipid homeostasis could potentially influence many essential biochemical pathways as cholesterol is required for synthesis of all steroid hormones and is critically involved in cell signaling, cell proliferation, phagocytosis, and cell motility.

In animal studies, we have demonstrated declines in male and female reproduction (Melin et al., 2014, 2015), and severe defects in neural development (Hrubec et al., 2018). We also have found that chronic oral dosing during breeding and gestation resulted in maternal absorption and transfer of C12 BAC and C16 BAC across the placenta to the fetus (Herron et al., 2019). BAC and DDAC also cross the blood-testis barrier to alter spermatogenesis (Melin, 2015). QACs disrupt cell signaling through alterations in sterol and oxysterol formation (Herron et al., 2019) and through receptor mediated G protein Gi/Go signaling (Higashijima et al., 1990). Taken together there is a wealth of evidence suggesting that BAC and DDAC have profound effects on basic biochemical and physiological processes. The data provided by this pilot study indicates that QACs may affect human responses as well.

## CONCLUSIONS

This study clearly demonstrated that QAC residues are present in human blood, and that alterations in inflammatory markers, mitochondrial function, and cholesterol synthesis were observed in a dose dependent manner with QAC concentration. Therefore, further studies are critically needed. It is not known if the exposure identified here correlates with disease in the human population. The primary sources of exposure and toxicokinetic properties such as absorption, distribution, metabolism, and elimination in humans is also not known. The limitations of our pilot study, including the small sample size and lack of information on other influential variables prevent us from answering these important questions. As the first study to identify possible systemic effects from chronic exposure in humans, however, the data clearly indicate the potential for health ramifications from QAC exposure.

## Data Availability

Data in the manuscript are available

---

**Supplemental Table 1.**
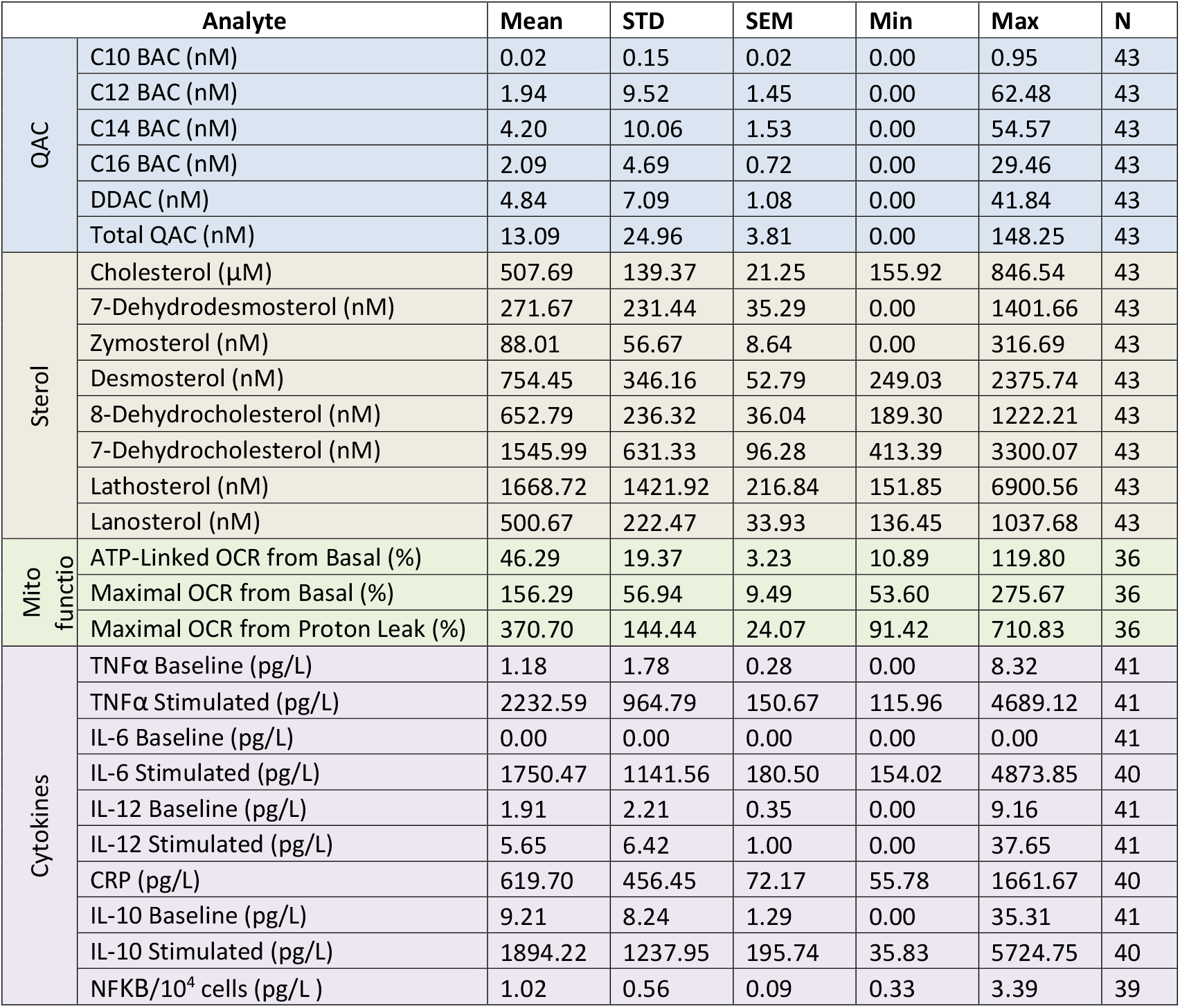
Descriptive statistics for all measured analytes. The varying sample number (N) reflects loss of some samples during the shipping and handling process.

**Supplemental Table 2.** Spearman’s Rank Correlations (r_s_) for C12 BAC. Blue cells designate r > 0.5 and p ≤ 0.05. Grey cells designate r > 0.5 and p ≤ 0.1. No notable correlations were observed above N=9 and are not included in the table.

| C12 |  |  |  |  |  |
| --- | --- | --- | --- | --- | --- |
| Analyte | N = 5 | N = 6 | N = 7 | N = 8 | N = 9 |
| Cholesterol - $r_2$ | 0.1000 | 0.3714 | 0.6071 | 0.7143 | 0.4167 |
| p | 0.7833 | 0.4194 | 0.1354 | 0.0480 | 0.2489 |
| 7-Dehydrodesmosterol - $r_2$ | 0.4000 | 0.2571 | 0.0000 | 0.0238 | -0.2833 |
| p | 0.4500 | 0.5639 | 1.0000 | 0.9309 | 0.4645 |
| Zymosterol - $r_2$ | -0.6000 | -0.4286 | 0.1071 | 0.2143 | 0.4500 |
| p | 0.3500 | 0.4194 | 0.7813 | 0.5796 | 0.2110 |
| Desmosterol - $r_2$ | -0.7000 | -0.2571 | 0.0000 | 0.1429 | 0.1667 |
| p | 0.2333 | 0.6583 | 1.0000 | 0.7059 | 0.6442 |
| 8-Dehydrocholesterol - $r_2$ | -0.7000 | -0.2571 | -0.0357 | -0.1667 | -0.1167 |
| p | 0.2333 | 0.6583 | 0.9684 | 0.7059 | 0.7757 |
| 7-Dehydrocholesterol - $r_2$ | -0.7000 | 0.2571 | 0.2143 | 0.2381 | 0.2333 |
| p | 0.2333 | 0.6583 | 0.6040 | 0.3595 | 0.5216 |
| Lathosterol - $r^2$ | 0.5000 | -0.1429 | -0.4286 | 0.0476 | 0.3167 |
| p | 0.3500 | 0.8028 | 0.3490 | 0.8850 | 0.3848 |
| Lanosterol - $r_2$ | 0.5000 | 0.7143 | 0.8214 | 0.8095 | 0.8333 |
| p | 0.3500 | 0.1028 | 0.0281 | 0.0188 | 0.0078 |
| ATP-Linked OCR - Basal - $r_2$ | -0.5000 | -0.7143 | -0.2143 | 0.0714 | -0.2000 |
| p | 0.4500 | 0.1361 | 0.6615 | 0.8394 | 0.6126 |
| Max. OCR - Basal - $r_2$ | 0.7000 | 0.3714 | 0.5357 | 0.6905 | 0.1833 |
| p | 0.1333 | 0.4194 | 0.1944 | 0.0583 | 0.6126 |
| Max. ORC - Proton Leak - $r_2$ | 0.5000 | 0.6571 | 0.6786 | 0.5000 | 0.3667 |
| p | 0.3500 | 0.1361 | 0.0886 | 0.1918 | 0.3128 |
| TNF-α Baseline - $r_2$ | 0.2236 | 0.3719 | 0.4454 | 0.0273 | 0.1584 |
| p | 0.6833 | 0.4194 | 0.3059 | 0.9309 | 0.6764 |
| TNF-α Stimulated - $r_2$ | -0.6000 | -0.2000 | -0.5000 | -0.4286 | -0.5667 |
| p | 0.3500 | 0.7139 | 0.2657 | 0.2967 | 0.1193 |
| IL-6 Stimulated - $r_2$ | 0.2000 | -0.1429 | -0.3214 | -0.0952 | -0.2333 |
| p | 0.6833 | 0.8028 | 0.4948 | 0.8394 | 0.5513 |
| IL-12 Baseline - $r_2$ | 0.3000 | 0.6000 | 0.7500 | 0.8333 | 0.4833 |
| p | 0.5167 | 0.1750 | 0.0532 | 0.0140 | 0.1768 |
| IL-12 Stimulated - $r_2$ | -0.9000 | -0.3714 | 0.1429 | -0.1667 | -0.3500 |
| p | 0.0833 | 0.4972 | 0.7207 | 0.7059 | 0.3599 |
| CRP - $r_2$ | 0.6000 | 0.6000 | 0.7500 | 0.1667 | 0.4167 |
| p | 0.2333 | 0.1750 | 0.0532 | 0.6629 | 0.2489 |
| IL-10 Baseline - $r_2$ | -0.3000 | -0.6000 | -0.5714 | -0.1905 | -0.3833 |
| p | 0.6833 | 0.2417 | 0.1944 | 0.6629 | 0.3128 |
| IL-10 Stimulated - $r_2$ | -0.6000 | -0.4286 | -0.3929 | 0.0714 | -0.2500 |
| p | 0.3500 | 0.4194 | 0.3950 | 0.8394 | 0.5216 |
| NFKB - $r_2$ | 0.5000 | 0.0857 | -0.1071 | -0.1905 | -0.2667 |
| p | 0.3500 | 0.8028 | 0.8430 | 0.6629 | 0.4927 |

**Supplemental Table 3.** Spearman’s Rank Correlations (r_s_) for C14 BAC. Blue cells designate r > 0.5 and p ≤ 0.05. Grey cells designate r > 0.5 and p ≤ 0.1. No notable correlations were observed above N=15 and are not included in the table.

| C14 |  |  |  |  |  |  |  |  |  |  |  |
| --- | --- | --- | --- | --- | --- | --- | --- | --- | --- | --- | --- |
| Analyte | N = 5 | N = 6 | N = 7 | N = 8 | N = 9 | N = 10 | N = 11 | N = 12 | N = 13 | N = 14 | N = 15 |
| Cholesterol - $r_2$ | -0.3000 | -0.0857 | 0.0000 | -0.1429 | 0.0000 | -0.2121 | -0.2727 | -0.3776 | -0.4451 | -0.3099 | -0.3750 |
| p | 0.6833 | 0.9194 | 0.9684 | 0.7498 | 0.9825 | 0.5599 | 0.4182 | 0.2274 | 0.1298 | 0.2806 | 0.1691 |
| 7-Dehydrodesmosterol - $r_2$ | -0.5000 | -0.2571 | -0.1429 | -0.4286 | -0.6000 | -0.6970 | -0.7727 | -0.8252 | -0.6868 | -0.5736 | -0.4893 |
| p | 0.4500 | 0.6583 | 0.7813 | 0.2967 | 0.0958 | 0.0311 | 0.0081 | 0.0017 | 0.0120 | 0.0350 | 0.0666 |
| Zymosterol - $r_2$ | -0.1000 | 0.3714 | 0.1429 | 0.2857 | 0.1000 | 0.2000 | 0.3545 | 0.1678 | 0.0330 | -0.0418 | 0.1214 |
| p | 0.9500 | 0.4194 | 0.7207 | 0.4631 | 0.7757 | 0.5599 | 0.2732 | 0.5883 | 0.9062 | 0.8915 | 0.6575 |
| Desmosterol - $r_2$ | 0.3000 | 0.6000 | 0.4286 | 0.5714 | 0.3000 | 0.1152 | 0.0818 | -0.0420 | -0.1319 | -0.1560 | 0.0286 |
| p | 0.5167 | 0.1750 | 0.3059 | 0.1302 | 0.4105 | 0.7329 | 0.7966 | 0.9037 | 0.6693 | 0.5944 | 0.9132 |
| 8-Dehydrocholesterol - $r_2$ | -0.6000 | -0.3143 | 0.1786 | 0.4524 | 0.3000 | -0.0545 | -0.0091 | -0.2378 | -0.4011 | -0.1473 | -0.3071 |
| p | 0.3500 | 0.5639 | 0.6615 | 0.2410 | 0.4105 | 0.8916 | 0.9892 | 0.4571 | 0.1758 | 0.6158 | 0.2649 |
| 7-Dehydrocholesterol - $r_2$ | -0.4000 | 0.0857 | 0.0714 | 0.0714 | 0.0667 | 0.0182 | -0.1091 | -0.2657 | -0.4231 | -0.1385 | -0.2929 |
| p | 0.5167 | 0.8028 | 0.8430 | 0.8394 | 0.8438 | 0.9457 | 0.7549 | 0.4040 | 0.1516 | 0.6375 | 0.2887 |
| Lathosterol - $r^2$ | -0.6000 | 0.0857 | -0.3214 | -0.3571 | -0.1167 | 0.0424 | 0.1545 | 0.0280 | -0.0604 | -0.0374 | -0.2179 |
| p | 0.3500 | 0.8028 | 0.4948 | 0.3921 | 0.7757 | 0.8916 | 0.6339 | 0.9212 | 0.8490 | 0.9035 | 0.4343 |
| Lanosterol - $r_2$ | -0.4000 | 0.2000 | 0.1429 | -0.1667 | -0.2333 | -0.1636 | -0.0727 | -0.2378 | -0.3516 | -0.3538 | -0.3286 |
| p | 0.5167 | 0.6583 | 0.7207 | 0.7059 | 0.5513 | 0.6567 | 0.8388 | 0.4571 | 0.2392 | 0.2149 | 0.2316 |
| ATP-Linked OCR - Basal - $r_2$ | 0.0000 | -0.3143 | -0.4286 | -0.4524 | -0.4500 | -0.1152 | -0.2273 | -0.3007 | -0.0824 | -0.1341 | -0.2107 |
| p | 1.0000 | 0.5639 | 0.3490 | 0.2680 | 0.2295 | 0.7588 | 0.5031 | 0.3425 | 0.7925 | 0.6485 | 0.4499 |
| Max. OCR - Basal - $r_2$ | 0.3000 | 0.1429 | 0.4643 | 0.1429 | -0.0667 | 0.2242 | 0.0091 | 0.0070 | 0.2033 | 0.1692 | 0.0143 |
| p | 0.5167 | 0.7139 | 0.2657 | 0.7059 | 0.8782 | 0.5139 | 0.9676 | 0.9737 | 0.4935 | 0.5526 | 0.9540 |
| Max. ORC - Proton Leak - $r_2$ | 0.3000 | 0.2571 | 0.5357 | 0.2619 | 0.1000 | 0.0061 | -0.0545 | -0.0909 | 0.0604 | 0.2484 | 0.1464 |
| p | 0.5167 | 0.5639 | 0.1944 | 0.5007 | 0.7757 | 0.9728 | 0.8815 | 0.7832 | 0.8348 | 0.3825 | 0.5934 |
| TNF-α Baseline - $r_2$ | -0.3536 | -0.1309 | 0.0000 | -0.4646 | -0.6337 | -0.7003 | -0.4738 | -0.6082 | -0.4197 | -0.5136 | -0.3586 |
| p | 0.6167 | 0.8028 | 0.9684 | 0.2680 | 0.0756 | 0.0275 | 0.1456 | 0.0400 | 0.1516 | 0.0616 | 0.1870 |
| TNF-α Stimulated - $r_2$ | 0.2000 | 0.5429 | 0.7143 | 0.8095 | 0.8167 | 0.6606 | 0.6818 | 0.4755 | 0.5274 | 0.6220 | 0.4429 |
| p | 0.6833 | 0.2417 | 0.0695 | 0.0188 | 0.0100 | 0.0394 | 0.0231 | 0.1154 | 0.0642 | 0.0192 | 0.0972 |
| IL-6 Stimulated - $r_2$ | -0.6000 | -0.6000 | -0.3214 | -0.1429 | -0.0333 | -0.0061 | 0.0000 | 0.0140 | 0.1099 | 0.1824 | 0.2929 |
| p | 0.3500 | 0.2417 | 0.4948 | 0.7498 | 0.9477 | 1.0000 | 0.9892 | 0.9562 | 0.7096 | 0.5221 | 0.2827 |
| IL-12 Baseline - $r_2$ | -0.3000 | -0.0857 | -0.2143 | -0.2857 | -0.4000 | -0.4667 | -0.5091 | -0.5594 | -0.5440 | -0.5868 | -0.6214 |
| p | 0.6833 | 0.9194 | 0.6615 | 0.5007 | 0.2906 | 0.1782 | 0.1140 | 0.0674 | 0.0581 | 0.0303 | 0.0156 |
| IL-12 Stimulated - $r_2$ | 0.7000 | 0.5429 | 0.6071 | 0.6429 | 0.1500 | 0.0545 | -0.1000 | -0.0280 | -0.2143 | -0.3495 | -0.4500 |
| p | 0.1333 | 0.2417 | 0.1354 | 0.0827 | 0.6764 | 0.8648 | 0.7757 | 0.9387 | 0.4819 | 0.2210 | 0.0944 |
| CRP - $r_2$ | -0.3000 | -0.1429 | -0.0714 | -0.0476 | 0.0500 | 0.3091 | 0.4727 | 0.5734 | 0.3956 | 0.5165 | 0.2786 |
| p | 0.6833 | 0.8028 | 0.9055 | 0.9309 | 0.8782 | 0.3677 | 0.1372 | 0.0521 | 0.1758 | 0.0591 | 0.3074 |
| IL-10 Baseline - $r_2$ | -0.3000 | -0.5429 | -0.7143 | -0.4048 | -0.2167 | -0.0909 | -0.2818 | -0.1608 | -0.3407 | -0.1209 | -0.0786 |
| p | 0.6833 | 0.2972 | 0.0886 | 0.3269 | 0.5816 | 0.8114 | 0.4021 | 0.6194 | 0.2550 | 0.6818 | 0.7827 |
| IL-10 Stimulated - $r_2$ | -0.9000 | -0.8857 | -0.7143 | -0.4524 | -0.1333 | -0.1702 | -0.3781 | -0.3187 | -0.2669 | 0.0132 | -0.1483 |
| p | 0.0833 | 0.0333 | 0.0886 | 0.2680 | 0.7421 | 0.6320 | 0.2484 | 0.3083 | 0.3733 | 0.9637 | 0.5934 |
| NFKB - $r_2$ | 0.1000 | 0.4286 | 0.0357 | 0.0714 | -0.2500 | -0.2485 | -0.3091 | -0.0979 | -0.0165 | 0.0154 | 0.0714 |
| p | 0.7833 | 0.3556 | 0.9055 | 0.8394 | 0.5216 | 0.4916 | 0.3560 | 0.7663 | 0.9639 | 0.9517 | 0.7926 |

**Supplemental Table 4.**
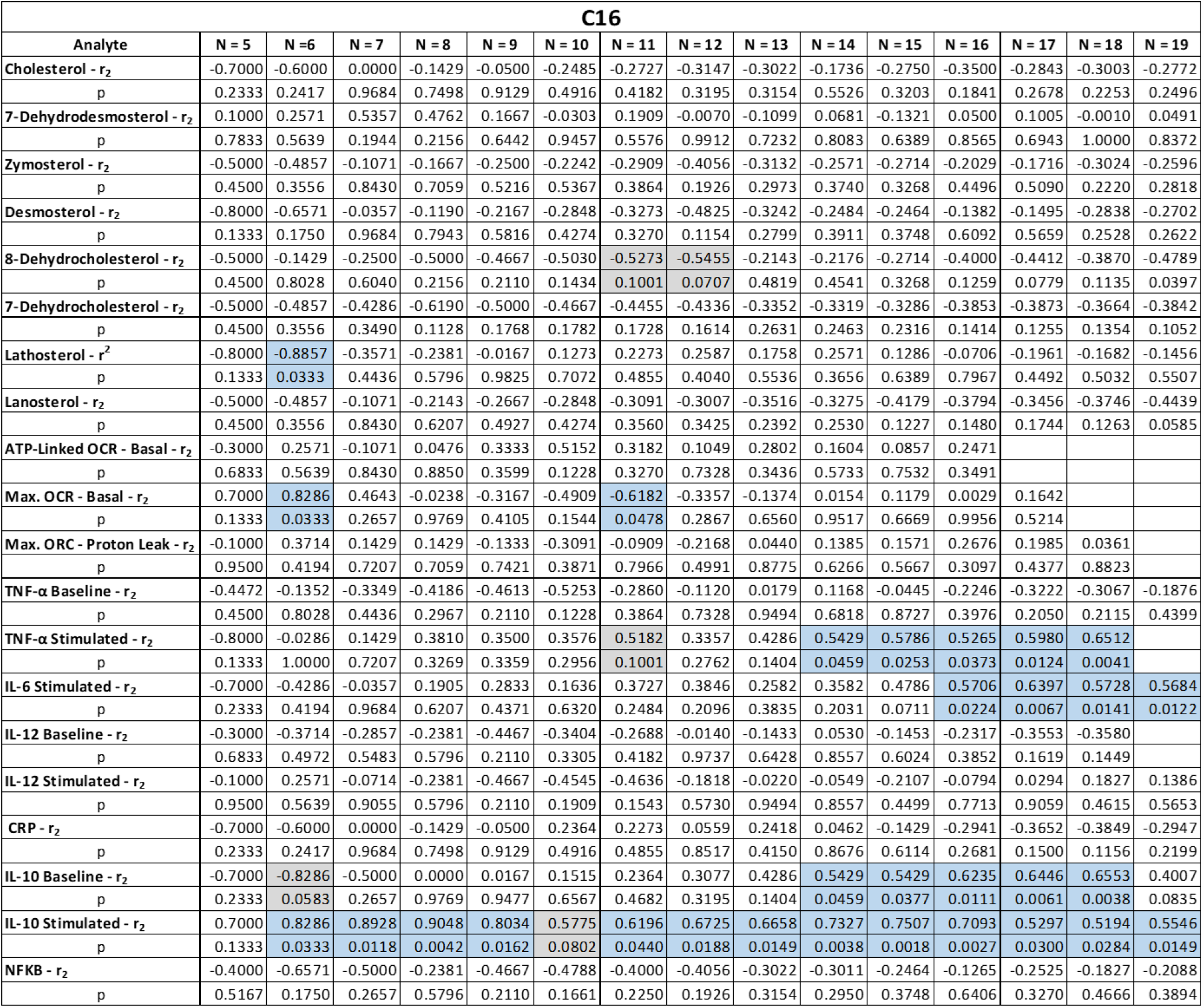
Spearman’s Rank Correlations (rs) for C16 BAC. Blue cells designate r > 0.5 and p ≤ 0.05. Grey cells designate r > 0.5 and p ≤ 0.1 Blank cells represents removed outliers, and samples lost during shipping. No notable correlations were observed above N=19 and are not included in the table.

**Supplemental Table 5.** Spearman’s Rank Correlations (r_s_) for DDAC. Blue cells designate r > 0.5 and p ≤ 0.05. Grey cells designate r > 0.5 and p ≤ 0.1. No notable correlations were observed above N=24 and are not included in the table.

| DDAC |  |  |  |  |  |  |  |  |  |  |  |  |  |  |  |  |  |  |  |  |
| --- | --- | --- | --- | --- | --- | --- | --- | --- | --- | --- | --- | --- | --- | --- | --- | --- | --- | --- | --- | --- |
| Analyte | N = 5 | N = 6 | N = 7 | N = 8 | N = 9 | N = 10 | N = 11 | N = 12 | N = 13 | N = 14 | N = 15 | N = 16 | N = 17 | N = 18 | N = 19 | N = 20 | N = 21 | N = 22 | N = 23 | N = 24 |
| Cholesterol - $r_2$ | 0.7000 | 0.8286 | 0.8929 | 0.8571 | 0.8000 | 0.4182 | 0.4364 | 0.2587 | 0.0714 | 0.0901 | 0.1536 | 0.2471 | 0.1838 | 0.2157 | 0.1316 | 0.0496 | -0.0429 | -0.1203 | 0.0198 | 0.0226 |
| p | 0.1333 | 0.0333 | 0.0118 | 0.0100 | 0.0126 | 0.2180 | 0.1728 | 0.4040 | 0.8065 | 0.7500 | 0.5756 | 0.3491 | 0.4727 | 0.3838 | 0.5852 | 0.8313 | 0.8545 | 0.5926 | 0.9263 | 0.9142 |
| 7-Dehydrodesmosterol - $r_2$ | 0.4000 | -0.2000 | 0.2500 | -0.1667 | 0.0333 | -0.2242 | -0.3364 | -0.0629 | -0.2637 | -0.3626 | -0.1929 | -0.0647 | -0.2108 | -0.1434 | -0.2614 | -0.1534 | -0.2338 | -0.1790 | -0.0395 | -0.1183 |
| p | 0.4500 | 0.7139 | 0.5483 | 0.7059 | 0.9129 | 0.5367 | 0.3130 | 0.8517 | 0.3835 | 0.2031 | 0.4901 | 0.8137 | 0.4152 | 0.5690 | 0.2785 | 0.5170 | 0.3063 | 0.4237 | 0.8584 | 0.5807 |
| Zymosterol - $r_2$ | 0.8000 | 0.8857 | 0.9286 | 0.7143 | 0.8000 | 0.7576 | 0.5909 | 0.5734 | 0.5714 | 0.3143 | 0.3357 | 0.2529 | 0.2966 | 0.3664 | 0.3877 | 0.3173 | 0.3351 | 0.4218 | 0.2441 | 0.2313 |
| p | 0.0833 | 0.0167 | 0.0064 | 0.0480 | 0.0126 | 0.0137 | 0.0562 | 0.0521 | 0.0425 | 0.2665 | 0.2160 | 0.3376 | 0.2430 | 0.1331 | 0.1002 | 0.1705 | 0.1362 | 0.0511 | 0.2585 | 0.2737 |
| Desmosterol - $r_2$ | 0.8000 | 0.8857 | 0.9286 | 0.7143 | 0.7667 | 0.4788 | 0.5545 | 0.4476 | 0.5000 | 0.4022 | 0.4107 | 0.4147 | 0.4534 | 0.3808 | 0.3842 | 0.3008 | 0.1234 | 0.2377 | 0.0830 | 0.0522 |
| p | 0.0833 | 0.0167 | 0.0064 | 0.0480 | 0.0191 | 0.1544 | 0.0759 | 0.1404 | 0.0812 | 0.1505 | 0.1261 | 0.1089 | 0.0677 | 0.1177 | 0.1035 | 0.1948 | 0.5890 | 0.2831 | 0.7025 | 0.8056 |
| 8-Dehydrocholesterol - $r_2$ | 0.0000 | 0.3714 | 0.5000 | 0.5238 | 0.5500 | 0.3818 | 0.2636 | 0.2587 | 0.1538 | 0.3231 | 0.3786 | 0.3353 | 0.2157 | 0.0836 | 0.1579 | 0.0451 | 0.0818 | 0.0412 | 0.1611 | 0.0217 |
| p | 1.0000 | 0.4194 | 0.2285 | 0.1697 | 0.1193 | 0.2629 | 0.4182 | 0.4040 | 0.6039 | 0.2530 | 0.1607 | 0.2000 | 0.3988 | 0.7357 | 0.5124 | 0.8462 | 0.7198 | 0.8522 | 0.4583 | 0.9174 |
| 7-Dehydrocholesterol - $r_2$ | 0.1000 | 0.4857 | 0.6429 | 0.6905 | 0.7333 | 0.6242 | 0.4455 | 0.3846 | 0.3297 | 0.3758 | 0.3107 | 0.2765 | 0.1691 | 0.0815 | 0.0842 | 0.0556 | 0.0610 | 0.0356 | 0.1561 | 0.0730 |
| p | 0.7833 | 0.2972 | 0.1105 | 0.0583 | 0.0274 | 0.0544 | 0.1634 | 0.2096 | 0.2631 | 0.1808 | 0.2535 | 0.2936 | 0.5090 | 0.7419 | 0.7264 | 0.8115 | 0.7886 | 0.8720 | 0.4723 | 0.7309 |
| Lathosterol - $r^2$ | -0.3000 | -0.5429 | 0.0357 | 0.1905 | 0.4333 | 0.4424 | 0.3455 | 0.3287 | 0.1923 | 0.1385 | 0.0964 | 0.2088 | 0.2941 | 0.1166 | 0.2018 | 0.2992 | 0.1221 | 0.2196 | 0.3172 | 0.2922 |
| p | 0.6833 | 0.2972 | 0.9055 | 0.6207 | 0.2295 | 0.1909 | 0.2861 | 0.2867 | 0.5172 | 0.6266 | 0.7241 | 0.4297 | 0.2470 | 0.6384 | 0.4017 | 0.1971 | 0.5929 | 0.3219 | 0.1390 | 0.1643 |
| Lanosterol - $r_2$ | 0.7000 | 0.8286 | 0.8929 | 0.7857 | 0.8500 | 0.7333 | 0.6364 | 0.3986 | 0.1648 | 0.0418 | -0.0500 | 0.0015 | 0.0969 | 0.1094 | 0.1781 | 0.2339 | 0.0656 | 0.0700 | 0.1863 | 0.0665 |
| p | 0.1333 | 0.0333 | 0.0118 | 0.0245 | 0.0058 | 0.0184 | 0.0370 | 0.1926 | 0.5785 | 0.8795 | 0.8626 | 0.9956 | 0.7084 | 0.6623 | 0.4619 | 0.3180 | 0.7755 | 0.7547 | 0.3918 | 0.7555 |
| ATP-Linked OCR - Basal - $r_2$ | -0.7000 | -0.2571 | -0.3214 | -0.0952 | 0.1000 | -0.0182 | 0.1273 | 0.0280 | 0.1703 | 0.0242 | 0.1571 | 0.3059 | 0.3750 | 0.2632 | 0.1561 | 0.0842 | 0.0260 | 0.0717 | 0.0198 | 0.0626 |
| p | 0.2333 | 0.6583 | 0.4948 | 0.8394 | 0.7757 | 0.9728 | 0.6935 | 0.9212 | 0.5660 | 0.9276 | 0.5667 | 0.2440 | 0.1360 | 0.5522 | 0.5171 | 0.7191 | 0.9079 | 0.7471 | 0.9263 | 0.7680 |
| Max. OCR - Basal - $r_2$ | -0.2000 | -0.3143 | -0.3214 | -0.5476 | -0.6833 | -0.7333 | -0.7455 | -0.3986 | -0.2198 | 0.0242 | 0.2036 | 0.2441 | 0.0956 | 0.1476 | 0.0737 | -0.0301 | 0.0338 | -0.0119 | -0.1354 | -0.2391 |
| p | 0.7833 | 0.5639 | 0.4948 | 0.1697 | 0.0509 | 0.0212 | 0.0119 | 0.2010 | 0.4704 | 0.9276 | 0.4578 | 0.3550 | 0.7084 | 0.5522 | 0.7592 | 0.9013 | 0.8811 | 0.9599 | 0.5364 | 0.2592 |
| Max. ORC - Proton Leak - $r_2$ | 0.9000 | 0.3714 | 0.4286 | 0.1190 | -0.0667 | -0.2121 | -0.2818 | -0.0909 | 0.0440 | 0.0593 | -0.0893 | -0.2059 | -0.2181 | -0.0547 | -0.0596 | -0.1188 | 0.0338 | -0.0333 | -0.0899 | -0.1722 |
| p | 0.0167 | 0.4194 | 0.3059 | 0.7498 | 0.8782 | 0.5599 | 0.4021 | 0.7832 | 0.8775 | 0.8319 | 0.7532 | 0.4430 | 0.3988 | 0.8306 | 0.8091 | 0.6169 | 0.8811 | 0.8839 | 0.6824 | 0.4194 |
| TNF-α Baseline - $r_2$ | 0.0513 | 0.3339 | -0.0371 | -0.2196 | 0.0348 | -0.1688 | 0.0286 | 0.1680 | 0.2688 | 0.1009 | 0.1956 | -0.0298 | -0.0530 | -0.1275 | -0.0241 | 0.0611 | 0.0316 | -0.0801 | -0.0836 | -0.1905 |
| p | 0.9500 | 0.4972 | 0.9684 | 0.6207 | 0.9129 | 0.6320 | 0.9244 | 0.5883 | 0.3633 | 0.7270 | 0.4738 | 0.9171 | 0.8389 | 0.6091 | 0.9225 | 0.7967 | 0.8900 | 0.7205 | 0.7025 | 0.3711 |
| TNF-α Stimulated - $r_2$ | -0.1000 | 0.0286 | 0.2857 | 0.4048 | 0.2833 | -0.0061 | 0.1182 | 0.2657 | 0.4231 | 0.2527 | 0.1679 | 0.2706 | 0.1054 | 0.1682 | 0.2930 | 0.3940 | 0.4000 | 0.4783 | 0.5069 | 0.5661 |
| p | 0.9500 | 0.9194 | 0.4948 | 0.2967 | 0.4371 | 1.0000 | 0.7138 | 0.3912 | 0.1459 | 0.3740 | 0.5406 | 0.3043 | 0.6803 | 0.4979 | 0.2199 | 0.0854 | 0.0725 | 0.0253 | 0.0144 | 0.0045 |
| IL-6 Stimulated - $r_2$ | 0.0000 | -0.1429 | -0.1429 | -0.1429 | -0.2167 | -0.2606 | -0.1000 | 0.1538 | 0.0495 | 0.0418 | 0.1071 | 0.2118 | 0.3431 | 0.2095 | 0.2491 | 0.2346 | 0.1935 | 0.1575 | 0.1266 | 0.2139 |
| p | 1.0000 | 0.8028 | 0.7813 | 0.7498 | 0.5816 | 0.4697 | 0.7757 | 0.6194 | 0.8632 | 0.8795 | 0.6953 | 0.4232 | 0.1744 | 0.3978 | 0.2989 | 0.3148 | 0.3957 | 0.4789 | 0.5638 | 0.3120 |
| IL-12 Baseline - $r_2$ | 0.6156 | 0.7827 | 0.2523 | 0.1437 | -0.2008 | -0.3222 | -0.4920 | -0.3643 | -0.4897 | -0.1914 | 0.0304 | -0.0692 | -0.0626 | -0.0930 | -0.0606 | -0.1543 | -0.1995 | -0.2814 | -0.1296 | -0.1550 |
| p | 0.2333 | 0.0583 | 0.5483 | 0.7059 | 0.6136 | 0.3677 | 0.1292 | 0.2463 | 0.0929 | 0.5121 | 0.9132 | 0.7967 | 0.8094 | 0.7109 | 0.8035 | 0.5128 | 0.3830 | 0.2033 | 0.5546 | 0.4685 |
| IL-12 Stimulated - $r_2$ | 0.3000 | 0.6000 | 0.1429 | -0.2381 | 0.0167 | 0.0545 | 0.1727 | 0.2727 | 0.3407 | 0.4374 | 0.4321 | 0.3441 | 0.3676 | 0.2590 | 0.0895 | 0.0737 | 0.0870 | 0.2061 | 0.0553 | 0.1679 |
| p | 0.5167 | 0.1750 | 0.7207 | 0.5796 | 0.9477 | 0.8648 | 0.5952 | 0.3787 | 0.2470 | 0.1158 | 0.1063 | 0.1880 | 0.1443 | 0.2941 | 0.7101 | 0.7528 | 0.7029 | 0.3531 | 0.7986 | 0.4290 |
| CRP - $r_2$ | 0.4000 | 0.6571 | 0.7857 | 0.6429 | 0.5333 | 0.6606 | 0.2455 | 0.0420 | 0.2308 | 0.1121 | 0.1464 | -0.0559 | -0.2132 | -0.0382 | 0.0158 | 0.1564 | 0.2234 | 0.2242 | 0.1482 | 0.1635 |
| p | 0.4500 | 0.1361 | 0.0395 | 0.0827 | 0.1323 | 0.0394 | 0.4512 | 0.8863 | 0.4367 | 0.6930 | 0.5934 | 0.8393 | 0.4097 | 0.8823 | 0.9454 | 0.5046 | 0.3260 | 0.3119 | 0.4951 | 0.4411 |
| IL-10 Baseline - $r_2$ | -0.1000 | -0.1429 | -0.1429 | -0.1429 | -0.2000 | 0.0545 | 0.2273 | 0.4056 | 0.4670 | 0.5077 | 0.5500 | 0.5588 | 0.3284 | 0.3870 | 0.1789 | 0.1053 | 0.2273 | 0.2829 | 0.3152 | 0.2365 |
| p | 0.3000 | 0.8028 | 0.7813 | 0.7498 | 0.6126 | 0.8648 | 0.4855 | 0.1845 | 0.1057 | 0.0641 | 0.0349 | 0.0258 | 0.1944 | 0.1115 | 0.4575 | 0.6534 | 0.3175 | 0.1996 | 0.1416 | 0.2628 |
| IL-10 Stimulated - $r_2$ | 0.5167 | 0.4286 | 0.5000 | 0.1667 | 0.1925 | 0.4134 | 0.4419 | 0.4518 | 0.4659 | 0.2288 | 0.3128 | 0.3782 | 0.1484 | 0.2313 | 0.2957 | 0.3813 | 0.4164 | 0.3050 | 0.3049 | 0.3544 |
| p | 0.1333 | 0.1333 | 0.3556 | 0.2285 | 0.6629 | 0.6126 | 0.2324 | 0.1728 | 0.1404 | 0.1103 | 0.4265 | 0.2535 | 0.1480 | 0.5659 | 0.3523 | 0.2171 | 0.0974 | 0.0613 | 0.1665 | 0.1563 |
| NFKB - $r_2$ | -0.7000 | -0.7000 | -0.7714 | -0.4286 | -0.6190 | -0.3500 | -0.3939 | -0.4636 | -0.5594 | -0.3626 | -0.3495 | -0.2429 | -0.2471 | -0.1103 | -0.1909 | -0.2561 | -0.3083 | -0.3857 | -0.2795 | -0.2638 |
| p | 0.2333 | 0.2333 | 0.1028 | 0.3490 | 0.1128 | 0.3599 | 0.2629 | 0.1543 | 0.0627 | 0.2240 | 0.2210 | 0.3820 | 0.3550 | 0.6734 | 0.4463 | 0.2886 | 0.1857 | 0.0851 | 0.2070 | 0.2229 |

**Supplemental Table 6.** Spearman’s Rank Correlations (r_s_) for Total QAC. Blue cells designate r > 0.5 and p ≤ 0.05. Grey cells designate r > 0.5 and p ≤ 0.1. No notable correlations were observed above N=26 and are not included in the table.

| Total QAC |  |  |  |  |  |  |  |  |  |  |  |  |  |  |  |  |  |  |  |  |  |  |
| --- | --- | --- | --- | --- | --- | --- | --- | --- | --- | --- | --- | --- | --- | --- | --- | --- | --- | --- | --- | --- | --- | --- |
| Analyte | N = 5 | N = 6 | N = 7 | N = 8 | N = 9 | N = 10 | N = 11 | N = 12 | N = 13 | N = 14 | N = 15 | N = 16 | N = 17 | N = 18 | N = 19 | N = 20 | N = 21 | N = 22 | N = 23 | N = 24 | N = 25 | N = 26 |
| Cholesterol - $r_2$ | 0.1000 | -0.0857 | 0.1786 | 0.2143 | 0.3500 | 0.4424 | 0.3818 | 0.3007 | 0.0824 | 0.2659 | 0.0821 | 0.1353 | 0.1740 | 0.0382 | 0.0860 | 0.0632 | 0.1069 | 0.0051 | -0.0583 | -0.1104 | -0.1062 | -0.1501 |
| p | 0.7833 | 0.9194 | 0.6615 | 0.5796 | 0.3359 | 0.1909 | 0.2365 | 0.3309 | 0.7785 | 0.3491 | 0.7630 | 0.6092 | 0.4967 | 0.8758 | 0.7209 | 0.7869 | 0.6491 | 0.9799 | 0.7916 | 0.6062 | 0.6123 | 0.4626 |
| 7-Dehydrodesmosterol - $r_2$ | 0.8000 | 0.8857 | 0.8929 | 0.4286 | 0.1333 | 0.3212 | 0.3364 | 0.3427 | 0.1429 | 0.3143 | 0.0679 | 0.1971 | 0.1275 | 0.1600 | 0.0246 | 0.0406 | -0.0494 | 0.0401 | -0.0603 | -0.1243 | -0.0685 | -0.0263 |
| p | 0.0833 | 0.0167 | 0.0118 | 0.2680 | 0.7090 | 0.3488 | 0.2994 | 0.2660 | 0.6298 | 0.2665 | 0.8025 | 0.4564 | 0.6187 | 0.5193 | 0.9168 | 0.8612 | 0.8324 | 0.8561 | 0.7846 | 0.5612 | 0.7446 | 0.8990 |
| Zymosterol - $r_2$ | -0.4000 | -0.2571 | 0.2143 | 0.1667 | 0.0000 | 0.0182 | -0.1545 | -0.1469 | -0.0824 | 0.1033 | -0.0571 | -0.1118 | -0.1520 | -0.0877 | -0.1316 | -0.1850 | -0.1987 | -0.1293 | -0.2095 | -0.3043 | -0.2677 | -0.2014 |
| p | 0.5167 | 0.6583 | 0.6040 | 0.6629 | 0.9825 | 0.9457 | 0.6535 | 0.6511 | 0.7925 | 0.7156 | 0.8425 | 0.6805 | 0.5594 | 0.7295 | 0.5902 | 0.4333 | 0.3862 | 0.5649 | 0.3358 | 0.1480 | 0.1951 | 0.3224 |
| Desmosterol - $r_2$ | 0.1000 | -0.0857 | 0.3214 | 0.5238 | 0.2500 | 0.3091 | -0.0182 | 0.0769 | -0.0385 | 0.1692 | 0.0750 | 0.2059 | 0.1152 | 0.2198 | 0.2526 | 0.1368 | 0.0675 | 0.1146 | -0.0128 | -0.1200 | -0.1331 | -0.2178 |
| p | 0.7833 | 0.9194 | 0.4436 | 0.1697 | 0.4927 | 0.3677 | 0.9676 | 0.8002 | 0.9062 | 0.5526 | 0.7827 | 0.4363 | 0.6526 | 0.3747 | 0.2920 | 0.5594 | 0.7669 | 0.6068 | 0.9550 | 0.5751 | 0.5244 | 0.2838 |
| 8-Dehydrocholesterol - $r_2$ | -0.1000 | -0.2000 | -0.2143 | 0.1905 | 0.0667 | -0.0667 | 0.1091 | -0.1469 | -0.2802 | -0.2440 | -0.3393 | -0.2147 | -0.2892 | -0.4014 | -0.4526 | -0.4917 | -0.3052 | -0.2626 | -0.3172 | -0.3626 | -0.4362 | -0.4174 |
| p | 0.9500 | 0.7139 | 0.6615 | 0.6207 | 0.8438 | 0.8648 | 0.7343 | 0.6511 | 0.3534 | 0.3998 | 0.2160 | 0.4232 | 0.2593 | 0.0998 | 0.0533 | 0.0292 | 0.1782 | 0.2368 | 0.1403 | 0.0823 | 0.0304 | 0.0349 |
| 7-Dehydrocholesterol - $r_2$ | -0.2000 | -0.2571 | 0.0714 | 0.2381 | 0.3167 | 0.2727 | 0.3455 | 0.0350 | 0.0440 | 0.1429 | 0.0214 | -0.0735 | -0.1495 | -0.2446 | -0.2895 | -0.3263 | -0.1455 | -0.1553 | -0.1937 | -0.2391 | -0.2785 | -0.2937 |
| p | 0.7833 | 0.6583 | 0.8430 | 0.5395 | 0.3848 | 0.4274 | 0.2861 | 0.9037 | 0.8775 | 0.6158 | 0.9336 | 0.7882 | 0.5659 | 0.3266 | 0.2286 | 0.1602 | 0.5277 | 0.4885 | 0.3742 | 0.2592 | 0.1772 | 0.1452 |
| Lathosterol - $r^2$ | -0.4000 | -0.0857 | 0.3214 | 0.3095 | 0.3500 | 0.4909 | 0.4182 | 0.3636 | 0.3846 | 0.4769 | 0.3286 | 0.2676 | 0.2181 | 0.0258 | -0.1000 | 0.0135 | 0.0338 | -0.0966 | 0.0247 | 0.0591 | 0.0731 | 0.1610 |
| p | 0.5167 | 0.9194 | 0.4436 | 0.4269 | 0.3359 | 0.1434 | 0.1926 | 0.2367 | 0.1888 | 0.0840 | 0.2264 | 0.3097 | 0.3934 | 0.9149 | 0.6834 | 0.9518 | 0.8811 | 0.6682 | 0.9083 | 0.7804 | 0.7251 | 0.4283 |
| Lanosterol - $r_2$ | -0.2000 | -0.2571 | 0.2143 | 0.0238 | -0.0333 | -0.0545 | 0.1000 | 0.0350 | 0.0275 | 0.1912 | 0.0143 | -0.0647 | -0.0576 | -0.0599 | -0.0606 | 0.0369 | 0.0032 | -0.1276 | -0.0341 | -0.0144 | -0.1081 | -0.0062 |
| p | 0.7833 | 0.6583 | 0.6040 | 0.9309 | 0.9477 | 0.8916 | 0.7549 | 0.9037 | 0.9206 | 0.5022 | 0.9540 | 0.8137 | 0.8241 | 0.8114 | 0.8035 | 0.8762 | 0.9887 | 0.5683 | 0.8762 | 0.9465 | 0.6045 | 0.9760 |
| ATP-Linked OCR - Basal - $r_2$ | -0.6000 | -0.4286 | -0.5000 | -0.3571 | -0.2000 | -0.0788 | -0.2364 | -0.2028 | -0.1758 | -0.2484 | -0.3000 | -0.3706 | -0.2181 | -0.0258 | -0.1035 | -0.1564 | -0.0532 | -0.1045 | -0.1462 | -0.1365 | -0.1077 | -0.1439 |
| p | 0.3500 | 0.4194 | 0.2657 | 0.3921 | 0.6126 | 0.8380 | 0.4855 | 0.5281 | 0.5660 | 0.3911 | 0.2767 | 0.1582 | 0.3988 | 0.9214 | 0.6728 | 0.5087 | 0.8192 | 0.6426 | 0.5039 | 0.5231 | 0.6071 | 0.4813 |
| Max. OCR - Basal - $r_2$ | -0.5000 | -0.4857 | -0.5357 | -0.6905 | -0.4000 | -0.5636 | -0.6364 | -0.7063 | -0.6538 | -0.7055 | -0.7036 | -0.5735 | -0.4681 | -0.2714 | -0.1123 | -0.0812 | -0.1338 | -0.1022 | -0.1670 | -0.2217 | -0.2915 | -0.3039 |
| p | 0.4500 | 0.3556 | 0.2285 | 0.0698 | 0.2906 | 0.0958 | 0.0404 | 0.0133 | 0.0183 | 0.0064 | 0.0046 | 0.0224 | 0.0600 | 0.2749 | 0.6465 | 0.7335 | 0.5618 | 0.4699 | 0.4446 | 0.2963 | 0.1571 | 0.1312 |
| Max. ORC - Proton Leak - $r_2$ | 0.2000 | 0.2571 | 0.2857 | 0.0000 | 0.1167 | -0.1879 | -0.1273 | -0.2168 | -0.2747 | -0.3143 | -0.3429 | -0.1059 | -0.0098 | 0.0857 | 0.0316 | 0.0857 | 0.0351 | -0.0638 | -0.0998 | -0.1461 | -0.1847 | -0.2185 |
| p | 0.6833 | 0.5639 | 0.4948 | 0.9769 | 0.7421 | 0.6076 | 0.7138 | 0.4991 | 0.3633 | 0.2735 | 0.2110 | 0.6968 | 0.9736 | 0.7295 | 0.0908 | 0.7143 | 0.8767 | 0.7779 | 0.6495 | 0.4941 | 0.3733 | 0.2823 |
| TNF-α Baseline - $r_2$ | -0.0513 | 0.2732 | 0.4335 | 0.0000 | 0.1826 | 0.3004 | 0.0347 | -0.1680 | -0.3034 | -0.1385 | -0.2522 | -0.1147 | -0.2264 | -0.3542 | -0.2226 | -0.1141 | -0.1829 | -0.2699 | -0.2516 | -0.1573 | -0.1625 | -0.2536 |
| p | 0.9500 | 0.5639 | 0.3059 | 0.9769 | 0.6126 | 0.3871 | 0.9029 | 0.6037 | 0.3154 | 0.6375 | 0.3607 | 0.6725 | 0.3774 | 0.1499 | 0.3576 | 0.6305 | 0.4252 | 0.2226 | 0.2450 | 0.4609 | 0.4365 | 0.2097 |
| TNF-α Stimulated - $r_2$ | 0.6000 | 0.6571 | 0.6786 | 0.7857 | 0.7500 | 0.6242 | 0.2182 | 0.2168 | 0.2143 | 0.3011 | 0.2107 | 0.0500 | 0.2083 | 0.2817 | 0.3404 | 0.4301 | 0.4312 | 0.4997 | 0.5553 | 0.5313 | 0.5423 | 0.4373 |
| p | 0.2333 | 0.1361 | 0.0886 | 0.0245 | 0.0230 | 0.0544 | 0.5031 | 0.4849 | 0.4704 | 0.2877 | 0.442 | 0.8479 | 0.4142 | 0.2528 | 0.1518 | 0.0588 | 0.0516 | 0.0188 | 0.0066 | 0.0083 | 0.0057 | 0.0263 |
| IL-6 Stimulated - $r_2$ | 0.7000 | 0.8286 | 0.4643 | 0.5238 | 0.5333 | 0.6121 | 0.4000 | 0.4336 | 0.3516 | 0.3802 | 0.3107 | 0.3529 | 0.3873 | 0.4592 | 0.5404 | 0.5113 | 0.5169 | 0.4692 | 0.4536 | 0.4122 | 0.3723 | 0.2923 |
| p | 0.1333 | 0.0333 | 0.2657 | 0.1697 | 0.1323 | 0.0602 | 0.2139 | 0.1542 | 0.2315 | 0.1755 | 0.2535 | 0.1764 | 0.1230 | 0.0557 | 0.0180 | 0.0223 | 0.0174 | 0.0285 | 0.0306 | 0.0459 | 0.0670 | 0.1461 |
| IL-12 Baseline - $r_2$ | -0.1000 | -0.4857 | -0.1786 | -0.0238 | 0.2833 | 0.3091 | 0.3182 | 0.0559 | -0.0165 | -0.1780 | -0.2000 | -0.2147 | -0.0221 | -0.1435 | -0.1474 | -0.1203 | -0.1494 | -0.2333 | -0.1779 | -0.2348 | -0.1016 | -0.1769 |
| p | 0.9500 | 0.3556 | 0.7207 | 0.9769 | 0.4371 | 0.3677 | 0.3270 | 0.8517 | 0.9639 | 0.5423 | 0.4738 | 0.4232 | 0.9360 | 0.5690 | 0.5458 | 0.6124 | 0.5166 | 0.2949 | 0.4151 | 0.2682 | 0.6279 | 0.3860 |
| IL-12 Stimulated - $r_2$ | -0.1000 | 0.0857 | 0.1429 | 0.3095 | -0.0833 | -0.1515 | -0.2909 | -0.3007 | -0.2857 | -0.3670 | -0.2500 | -0.1853 | -0.1348 | -0.2528 | -0.1684 | -0.2677 | -0.2416 | -0.3213 | -0.1561 | -0.2574 | -0.2138 | -0.08 |
| p | 0.9500 | 0.8028 | 0.7207 | 0.4269 | 0.8438 | 0.6818 | 0.3864 | 0.3425 | 0.3436 | 0.1973 | 0.3677 | 0.4909 | 0.6053 | 0.3101 | 0.4891 | 0.2528 | 0.2901 | 0.1448 | 0.4751 | 0.2237 | 0.3032 | 0.6956 |
| CRP - $r_2$ | 0.0000 | 0.2000 | 0.1429 | 0.2857 | -0.1000 | 0.2000 | 0.1636 | 0.3566 | 0.4890 | 0.5824 | 0.5750 | 0.3176 | 0.1201 | -0.0568 | 0.1018 | 0.2286 | 0.2597 | 0.1903 | 0.2194 | 0.2643 | 0.2708 | 0.1911 |
| p | 1.0000 | 0.6583 | 0.7207 | 0.4631 | 0.8096 | 0.5599 | 0.6144 | 0.2463 | 0.0889 | 0.0303 | 0.0264 | 0.2257 | 0.6390 | 0.8242 | 0.6728 | 0.3276 | 0.2520 | 0.3917 | 0.3108 | 0.2096 | 0.1886 | 0.3463 |
| IL-10 Baseline - $r_2$ | 0.4000 | 0.6571 | 0.5357 | 0.6429 | 0.6167 | 0.6727 | 0.5273 | 0.5944 | 0.5934 | 0.5429 | 0.5679 | 0.5853 | 0.6275 | 0.6202 | 0.3772 | 0.1805 | 0.2416 | 0.2908 | 0.3231 | 0.273 | 0.3569 | 0.3039 |
| p | 0.4500 | 0.1361 | 0.1944 | 0.0827 | 0.0756 | 0.0351 | 0.0936 | 0.0428 | 0.0340 | 0.0459 | 0.0286 | 0.0186 | 0.0081 | 0.0070 | 0.1104 | 0.4409 | 0.2875 | 0.1869 | 0.1315 | 0.1946 | 0.0798 | 0.1303 |
| IL-10 Stimulated - $r_2$ | 1.0000 | 0.6571 | 0.7857 | 0.8571 | 0.9000 | 0.8909 | 0.9182 | 0.7566 | 0.6549 | 0.5743 | 0.3753 | 0.4857 | 0.5714 | 0.6185 | 0.5529 | 0.5897 | 0.6171 | 0.4999 | 0.4705 | 0.5210 | 0.5432 | 0.5693 |
| p | 0.0000 | 0.1361 | 0.0395 | 0.0100 | 0.0018 | 0.0009 | 0.0000 | 0.0059 | 0.0171 | 0.0334 | 0.1649 | 0.0572 | 0.0179 | 0.0072 | 0.0152 | 0.0071 | 0.0034 | 0.0188 | 0.0244 | 0.0098 | 0.0056 | 0.0028 |
| NFKB - $r_2$ | 0.0000 | -0.4286 | -0.0714 | 0.0476 | -0.2667 | -0.1394 | 0.0273 | 0.1818 | 0.1593 | 0.2220 | 0.2821 | 0.3059 | 0.2059 | 0.2301 | 0.3035 | 0.2632 | 0.1481 | 0.1971 | 0.1719 | 0.0800 | -0.0323 | -0.0168 |
| p | 1.0000 | 0.4194 | 0.9055 | 0.8850 | 0.4927 | 0.7072 | 0.9244 | 0.5578 | 0.5912 | 0.4356 | 0.3011 | 0.2440 | 0.4208 | 0.3523 | 0.2033 | 0.2584 | 0.5166 | 0.3749 | 0.4283 | 0.7065 | 0.8786 | 0.9361 |

